# A Myasthenia Gravis genomewide association study of three cohorts identifies Agrin as a novel risk locus

**DOI:** 10.1101/2020.10.26.20219261

**Authors:** Apostolia Topaloudi, Zoi Zagoriti, Alyssa C. Flint, Melanie B. Martinez, Zhiyu Yang, Fotis Tsetsos, Yiolanda-Panayiota Christou, George Lagoumintzis, Evangelia Yannaki, Eleni Papanicolaou-Zamba, Konstantinos Poulas, John Tzartos, Xanthippi Tsekmekidou, Kalliopi Kotsa, Efstratios Maltezos, Nikolaos Papanas, Dimitrios Papazoglou, Ploumis Passadakis, Athanasios Roumeliotis, Stefanos Roumeliotis, Marios Theodoridis, Elias Thodis, Stylianos Panagoutsos, John Yovos, John A. Stamatoyannopoulos, Kleopas A. Kleopa, Socrates Tzartos, Marianthi Georgitsi, Peristera Paschou

**Affiliations:** Department of Biological Sciences, Purdue University, West Lafayette, IN, United States; Department of Pharmacy, University of Patras, Rio, Greece; Department of Molecular Biology and Genetics, Democritus University of Thrace, Alexandroupoli, Greece; The Cyprus Institute of Neurology and Genetics, Nicosia, Cyprus; Department of Hematology, George Papanicolaou Hospital, Thessaloniki, Greece; Department of Neuroepidemiology and Centre for Neuromuscular Disorders, The Cyprus Institute of Neurology and Genetics and Cyprus School of Molecular Medicine, Nicosia, Cyprus; Tzartos NeuroDiagnostics, Athens, Greece; Division of Endocrinology and Metabolism-Diabetes Center, 1st Department of Internal Medicine, AHEPA University Hospital, Aristotle University of Thessaloniki, Thessaloniki, Greece; Diabetes Center, 2nd Department of Internal Medicine, Alexandroupolis University General Hospital, Democritus University of Thrace, Alexandroupoli, Greece; Department of Nephrology, Alexandroupolis University General Hospital, Democritus University of Thrace, Alexandroupoli, Greece; Division of Nephrology and Hypertension, 1st Department of Internal Medicine, AHEPA University Hospital, Aristotle University of Thessaloniki, Thessaloniki, Greece; Departments of Medicine and Genome Sciences, University of Washington, Seattle, WA, United States; Department of Neuroscience and Centre for Neuromuscular Disorders, The Cyprus Institute of Neurology and Genetics and Cyprus School of Molecular Medicine, Nicosia, Cyprus; Hellenic Pasteur Institute, Athens, Greece; 1st Laboratory of Medical Biology-Genetics, Department of Medicine, Aristotle University of Thessaloniki, Thessaloniki, Greece

**Author notes:** **Corresponding authors:** Dr. Peristera Paschou, Lilly Hall of Life Sciences, 915 W. State Street, Room 1-225, West Lafayette, IN 47907., Dr. Marianthi Georgitsi, Department of Medicine, Sector of Biological Sciences and Preventive Medicine Aristotle University of Thessaloniki, University Campus, 54124 Thessaloniki, Greece. equal contribution senior authors (Marianthi Georgitsi, Peristera Paschou).

## Abstract

**Background:** Myasthenia Gravis (MG) is a rare autoimmune disorder affecting the neuromuscular junction. Here, we investigate the genetic architecture of MG performing a genomewide association study (GWAS) of the largest MG dataset analyzed to date.

**Methods:** We integrated GWAS from three different datasets (1,401 cases, 3,508 controls) and performed MG GWAS and onset-specific analyses. We also carried out HLA fine-mapping, gene-based, gene ontology and tissue enrichment analyses and investigated genetic correlation to other autoimmune disorders.

**Findings:** We observed the strongest MG association to *TNFRSF11A* (rs4369774, p=1.09×10^−13^; OR=1.4). Gene-based analysis revealed *AGRN* as a novel MG susceptibility gene. HLA fine-mapping pointed to two independent loci significantly associated with MG: *HLA-DRB1* (with a protective role) and *HLA-B*. MG onset-specific analysis, reveals differences in the genetic architecture of Early-Onset vs Late-Onset MG. Furthermore, we find MG to be genetically correlated with Type 1 Diabetes, Rheumatoid Arthritis and late-onset Vitiligo.

**Interpretation:** Overall, our results are consistent with previous studies highlighting the role of the HLA and *TNFRSF11A* in MG etiology and different risk genes in EOMG vs LOMG. Furthermore, our gene-based analysis implicates, for the first time, *AGRN* as a MG susceptibility locus. *AGRN* encodes agrin, which is involved in neuromuscular junction formation. Mutations in *AGRN* have been found to underlie congenital myasthenic syndrome. Gene ontology analysis suggests an intriguing role for symbiotic processes in MG etiology. We also uncover genetic correlation of MG to Type 1 Diabetes, Rheumatoid Arthritis and late-onset Vitiligo, pointing to shared underlying genetic mechanisms.

**Funding:** This work was supported by NSF award #1715202, the European Social Fund and Greek funds through the National Strategic Reference Framework (NSRF) THALES Programme 2012–2015 and the NSRF ARISTEIA II Programme 2007–2013 to PP, and grants from the Association Francaise contre les Myopathies (AFM, Grant No. 80077) to ST.

**Research in context:** *Evidence before this study:* Myasthenia Gravis (MG) is a complex disease caused by the interaction of genetic and environmental factors that lead to autoimmune activation. Previous studies have shown that the human leukocyte antigen (HLA) displays the most robust genetic association signals to MG. Additional susceptibility genes that have emerged through genomewide association studies (GWAS), include *CTLA4* and *TNFRSF11A*. Previous studies also support the hypothesis of distinct risk loci underlying Early-Onset versus Late-Onset MG subgroups (EOMG vs LOMG). For instance, *PTPN22* and *TNIP1* genes have been implicated in EOMG and *ZBTB10* in LOMG. In the GWAS studies published so far, *HLA* and *TNFRSF11A* associations appear to be confirmed; however, the association of other implicated genes still requires replication.

*Added value of this study:* We present the largest GWAS for MG to date, integrating three different datasets. We identify *AGRN* as a novel MG risk locus and replicate previously reported susceptibility loci, including HLA, *TNFRSF11A, and CTLA4*. Our analysis also supports the existence of a different genetic architecture in EOMG vs LOMG and identifies a region between *SRCAP* and *FBRS* as a novel EOMG risk locus. Additionally, through HLA fine-mapping, we observe different HLA genes implicated in EOMG vs LOMG (*HLA-B* and *HLA-DRB1* respectively). Finally, we detect positive genetic correlation of MG with other autoimmune disorders including Type 1 Diabetes, Rheumatoid Arthritis, and late-onset Vitiligo, suggesting a shared genetic basis across them.

*Implications of all the available evidence:* Our study sheds light into the etiology of MG identifying *AGRN* as a novel risk locus. *AGRN* encodes agrin, a protein with a significant role in the formation of the neuromuscular junction and mutations in this gene have been associated with congenital myasthenic syndrome. Our findings hint to an intriguing hypothesis of symbiotic processes underlying MG pathogenesis and points to muscle growth and development in EOMG and steroid hormones synthesis in LOMG. The observed genetic correlations between MG and certain other autoimmune disorders could possibly underlie comorbidity patterns across this group of disorders.

## Introduction

Myasthenia gravis (MG) is a rare autoimmune disorder mediated by autoantibodies that bind to specific proteins at the postsynaptic membrane of the neuromuscular junction (NMJ), thus attenuating endplate potential and neuromuscular transmission. Muscle fatigue constitutes the typical feature of the disease and common clinical manifestations include ptosis, diplopia, bulbar and limb muscle weakness.^1^ MG cases are classified into three distinct subgroups based on antibody specificity: Pathogenic autoantibodies against the skeletal muscle acetylcholine receptor (AChR) are detected in approximately 85% of MG cases, while in the remaining patients, autoantibodies are directed against the muscle-specific receptor tyrosine kinase (MuSK) or the lipoprotein-receptor-related protein 4 (LRP4).^2^ Recently, an additional group of MG-related autoantibodies targeting agrin, an LRP4 interaction partner that mediates the activation of MuSK and subsequently the formation of AChR clusters in the postsynaptic membrane,^3^ was identified in AChR/MuSK/LRP4 seropositive and seronegative MG patients.^4^ Based on disease onset age, anti-AChR MG patients can be further divided into two subtypes characterized by sex bias; in early-onset (EOMG; <50 y.o), a female predominance is evident, whereas in late-onset (LOMG; ≥50 y.o and particularly at ≥60 y.o), males are mostly affected.^2,5^ This immunological and epidemiological heterogeneity is suggestive of discrete pathogenetic mechanisms among the different MG subgroups.

MG is a complex disease, caused by the interaction of genetic and environmental factors that contribute to autoimmune activation.^6^ So far, the human leukocyte antigen (HLA) region displays the most robust genetic association signals to MG. ^7,8^ Additional non-HLA susceptibility genes have also emerged through genomewide association studies (GWAS),^9–11^ including *CTLA4* and *TNFRSF11A*. Previous studies also support the hypothesis of distinct risk loci underlying EOMG compared to LOMG.^11^ For instance, *PTPN22* and *TNIP1* genes have been implicated in EOMG^9^ and *ZBTB10* in LOMG.^11^ In GWAS studies published so far, *HLA* and *TNFRSF11A* associations appear to be confirmed;^8,11^ however, the association of other implicated genes still requires replication.

Considering the rarity of the disorder and the diversity of MG sub-phenotypes,^5^ unravelling the genetic architecture of MG has proven challenging, partly owing to the relatively small sample sizes available even in multi-centered studies. These factors render the identification of common risk variants with small or modest effect sizes a difficult task. Here, we present the results of a GWAS meta-analysis of a total of 1,401 cases and 3,508 controls, representing the largest MG sample size analyzed to date.

## Methods

### Study design

We performed a MG GWAS meta-analysis using genotype data from three different sources: the final meta-analysis dataset included 1,401 MG cases and 3,508 controls: 1) 196 Greek and Greek-Cypriot cases and 1,057 ancestry-matched controls (novel dataset), 2) 964 European-American cases and 1,985 ancestry-matched controls, available through dbGaP, and 3) 241 cases and 466 controls, available through the UK biobank. A detailed description of clinical inclusion criteria for each study is provided in supplementary text. Stringent quality control procedures and genotype imputation are described in supplementary text. The inverse-variance method as implemented in METAL^12^ was used for the meta-analysis. Heterogeneity was assessed with Cochran’s I^2^ statistic; only SNPs present in all three studies, with MAF >0·01 in each study and heterogeneity p-value >0·05, were included.

### Heritability estimation

MG heritability was calculated via the GCTA-GREML^13^ method on the combined imputed datasets, assuming a population prevalence of 0·02%^1^.

### Gene-Based and Tissue Specificity Analysis

Gene-based analyses were performed with MAGMA^14^ v1.08; p-values were adjusted with Bonferroni correction. The extended HLA region (hg19, chr6 25-33 Mb) was excluded from gene-based analyses. FUMA’s SNP2GENE^15^ tissue specificity analyses were performed to test whether there is any association between tissue-specific gene expression profiles using GTEx v.8 RNA-seq data and disease-genes. ClusterProfiler^16^ was used to perform enrichment tests of the top 100 and 200 genes from the gene-based analysis for Biological Process GO clusters.

### Genetic correlation with autoimmune disorders

Genetic correlation analysis of MG to other autoimmune disorders was performed through LDSC.^17^ Publicly available summary statistics from 12 autoimmune disorders GWAS^18–25^ for European ancestry samples were used (Table S1). Only SNPs that were included in the HapMap3 reference were included in the analyses. Pre-calculated LD scores from 1000Genomes European data were used. All studies included had a heritability z-score >4.

### HLA imputation and association tests

Imputation of HLA antigens was performed in each of the three datasets using SNP2HLA;^26^ Type 1 Diabetes Genetics Consortium data served as the reference panel. Imputed variants with info score >0·9 in each dataset were merged in one megaset. For each variant type (SNPs, HLA allele and HLA amino acid), association tests were performed, followed by conditional regression, in an effort to identify independent HLA associations if the conditional p-value was <5×10^−8^. All analyses were controlled for PCs suggested by SMARTPCA. For the significantly associated loci, reciprocal analyses were conducted to examine if the association signal can be attributed to the amino acid polymorphism.

### Onset-specific analyses

Following imputation, analyses were performed for EOMG vs LOMG datasets. The EOMG group consisted of 455 cases with age of onset less than 50 years old (66 in Greeks/Greek-Cypriots, 322 in dbGaP, and 67 in UK Biobank), and the LOMG group consisted of 946 cases with age of onset at 50 years old and above (130 in Greeks/Greek-Cypriots, 642 in dbGaP, and 174 in UK Biobank). This age cut-off has been previously proposed to serve as a useful criterion for the bimodal distribution of ages of onset in MG genetic studies, without significant deviation in results.^8,11^

## Results

### Genome-Wide Association Study

First, we performed GWAS on each independent dataset (Figure S1 and S2), followed by a GWAS meta-analysis across the three datasets (5,755,778 SNPs in 1,401 MG cases and 3,508 controls) (Figure 1A). The meta-analysis genomic control factor λ (λ_GC_=1·035) did not show evidence for residual stratification. The LD score reported intercept was 1·01 (SE=0·01), suggesting successful control of ancestry effects. The MG SNP-heritability calculated by GREML was h^2^_SNP_= 0·37 (SE=0·05, P=1·39×10^−15^).

**Figure 1:**
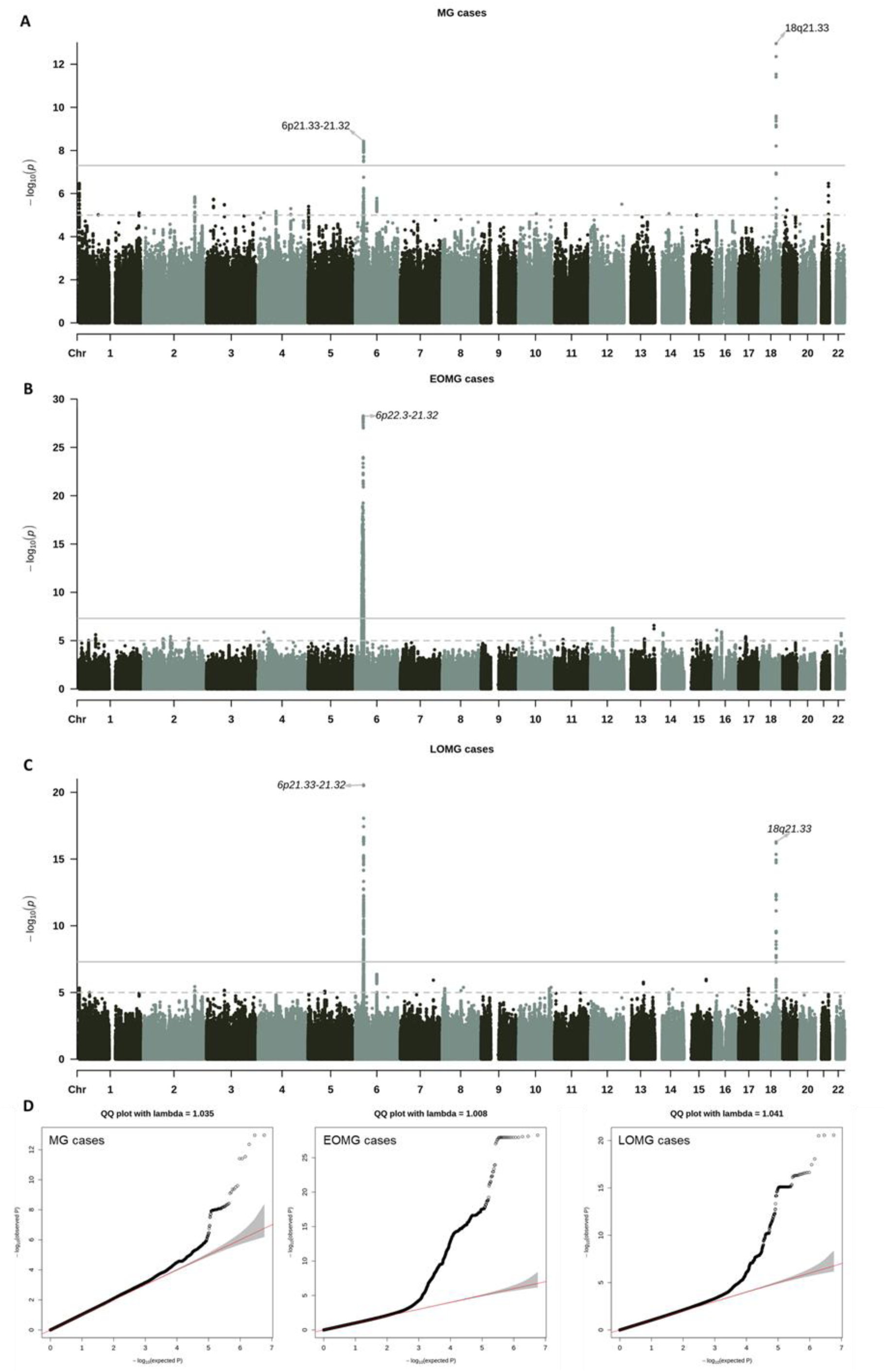
Manhattan plot and quartile-quartile plots for MG GWAS meta-analysis. A) GWAS meta-analysis of 1,401 MG cases and 3,508 controls, B) GWAS meta-analysis of 455 EOMG cases and 3,508 controls, C) GWAS meta-analysis of 946 LOMG cases and 3,508 controls. The solid horizontal line indicates the p-value threshold for the genomewide significance (5×10^−8^), whereas the dashed line indicates the p-value threshold of 10^−5^. Genome-wide significant risk loci are annotated. D) Quartile-quartile plots showing the distribution of expected vs observed p values of the respective GWAS meta-analyses. The 95% confidence interval of expected values is indicated in gray.

Table 1 shows results of our meta-analysis compared to previous studies. Our top SNP was rs4369774 (p=1·09×10^−13^, OR=1·4, 95% CI 1·29-3·62). It is located in an intron of the *Tumor Necrosis Factor Receptor Superfamily Member 11A* (*TNFRSF11A*) gene (Figure S3A). We identified an additional significantly associated locus in the *HLA-DQA1* gene with top SNP rs34481484 (p=3·72×10^−9^; OR=2·11, 95% CI 1·65-5·19). Overall, we identified 24 LD-independent regions with p<10^−5^ for index SNPs and the variants within 3-Mb windows and r^2^>0·1 with index SNPs (Table S2).

**Table 1:**
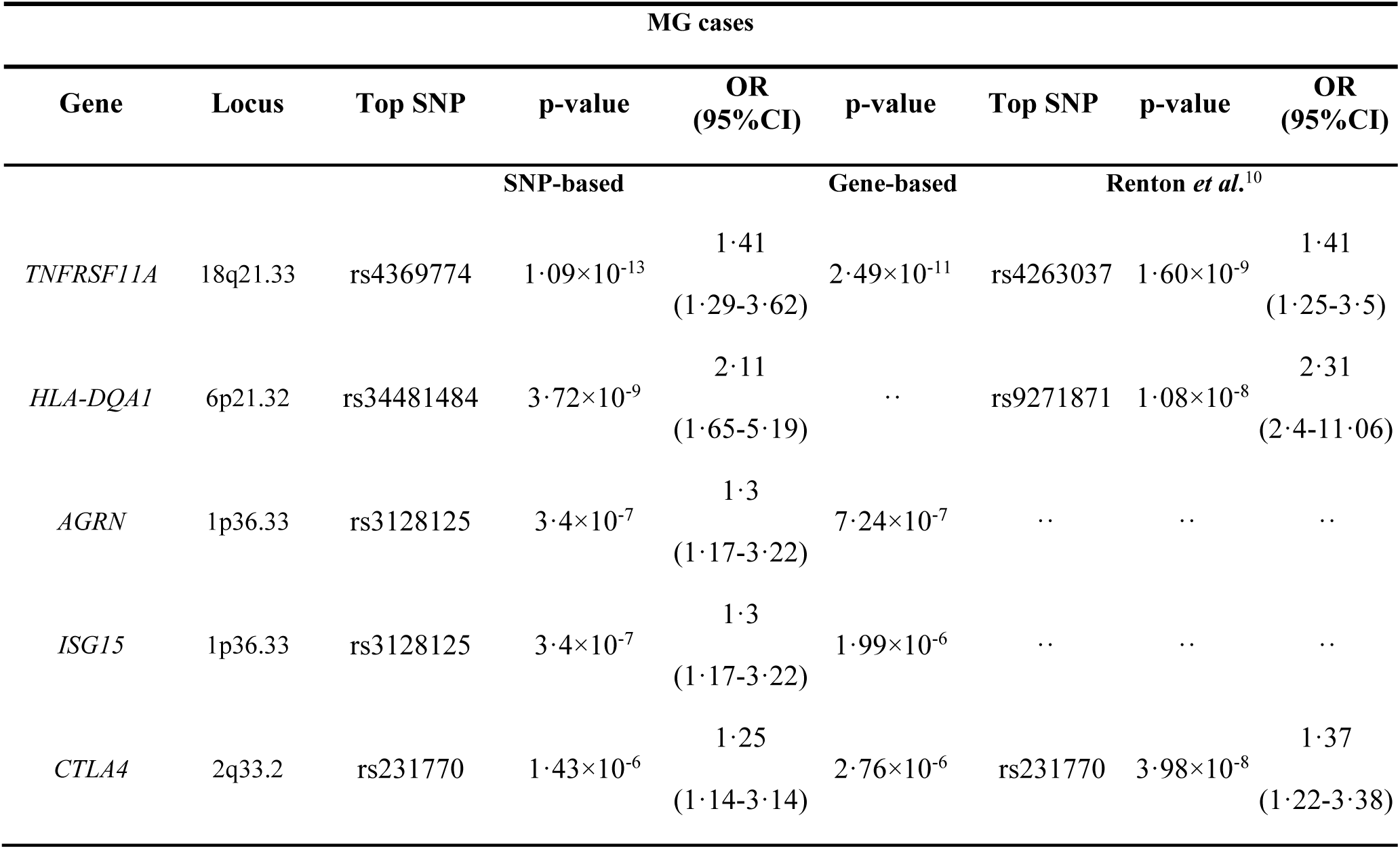
Genome-wide significant loci in MG. Genetic loci detected as significantly associated with MG in comparison to previous studies. For the non-HLA loci, SNP column corresponds to the most significant SNP nearest to each gene within 10-kb. The significance threshold for gene-based analysis is p=2·78×10^−6^. For the HLA region (6p22.1-21.3), we report the top SNP in each study and the closest gene to the SNP.

We then performed gene-based association tests with the extended HLA region (hg19, chr6 25-33 Mb) excluded from analysis due to the complicated LD pattern. The HLA region was analyzed in detail via HLA fine-mapping as described later in this manuscript. With all MG cases analyzed jointly, four genes, namely *TNFRSF11A, CTLA4, AGRN*, and *ISG15*, were found significantly associated with MG after correcting for 17,994 gene tests (p=2·78×10^−6^) (Figure S4A). *ISG15* is located only 5.58-kb upstream of *AGRN* on chromosome 1q36.33. Therefore, we performed conditional tests using the genotyped data to investigate the independent variants in the region (Supplementary material). The only independent SNP (rs3128125) was located in *AGRN* (Figure S5).

We explored GO-term-based networks among the top 100 and top 200 genes from our gene-based analysis (excluding HLA region) (Figure 2Α, S6A). Although not reaching genomewide statistical significance, gene clusters related to transcription regulation and symbiotic processes emerged at the top. Tissue specificity analysis in 54 and 30 general human tissues from GTEx v.8, did not identify statistically significant tissue-specific enrichment of gene expression of our top hits using the Bonferroni correction (30 tissues: p<1·67×10^−3^; 54 tissues: p<9·26×10^−4^) (Table 2 and Figures S7A and 8A). Brain cerebellum, subcutaneous adipose, thyroid, and skeletal muscle tissues emerged as the top tissues enriched in MG-associated genes.

**Table 2:** Tissue-specific enrichment analysis of genes associated with MG. The top five tissues from each tissue-specificity analysis are presented. Enrichment was tested in 54 general human tissue types from 948 donors using GTEx v.8 RNA-seq data. The significance threshold for the tissue-specific test was calculated using the Bonferroni correction for 54 tests (p=9·26×10^−4^).

| MG |  | EOMG |  | LOMG |  |
| --- | --- | --- | --- | --- | --- |
| Tissue | p-value | Tissue | p-value | Tissue | p-value |
| Brain Cerebellum | 0.02 | Colon Sigmoid | 0.01 | Thyroid | 0.02 |
| Brain Cerebellar Hemisphere | 0.03 | Brain Hypothalamus | 0.02 | Brain Cerebellum | 0.03 |
| Adipose Subcutaneous | 0.09 | Esophagus Muscularis | 0.05 | Pituitary | 0.04 |
| Thyroid | 0.10 | Brain Caudate basal ganglia | 0.07 | Brain Cerebellar Hemisphere | 0.04 |
| Muscle Skeletal | 0.11 | Esophagus Gastroesophageal Junction | 0.08 | Kidney Medulla | 0.12 |

**Figure 2:**
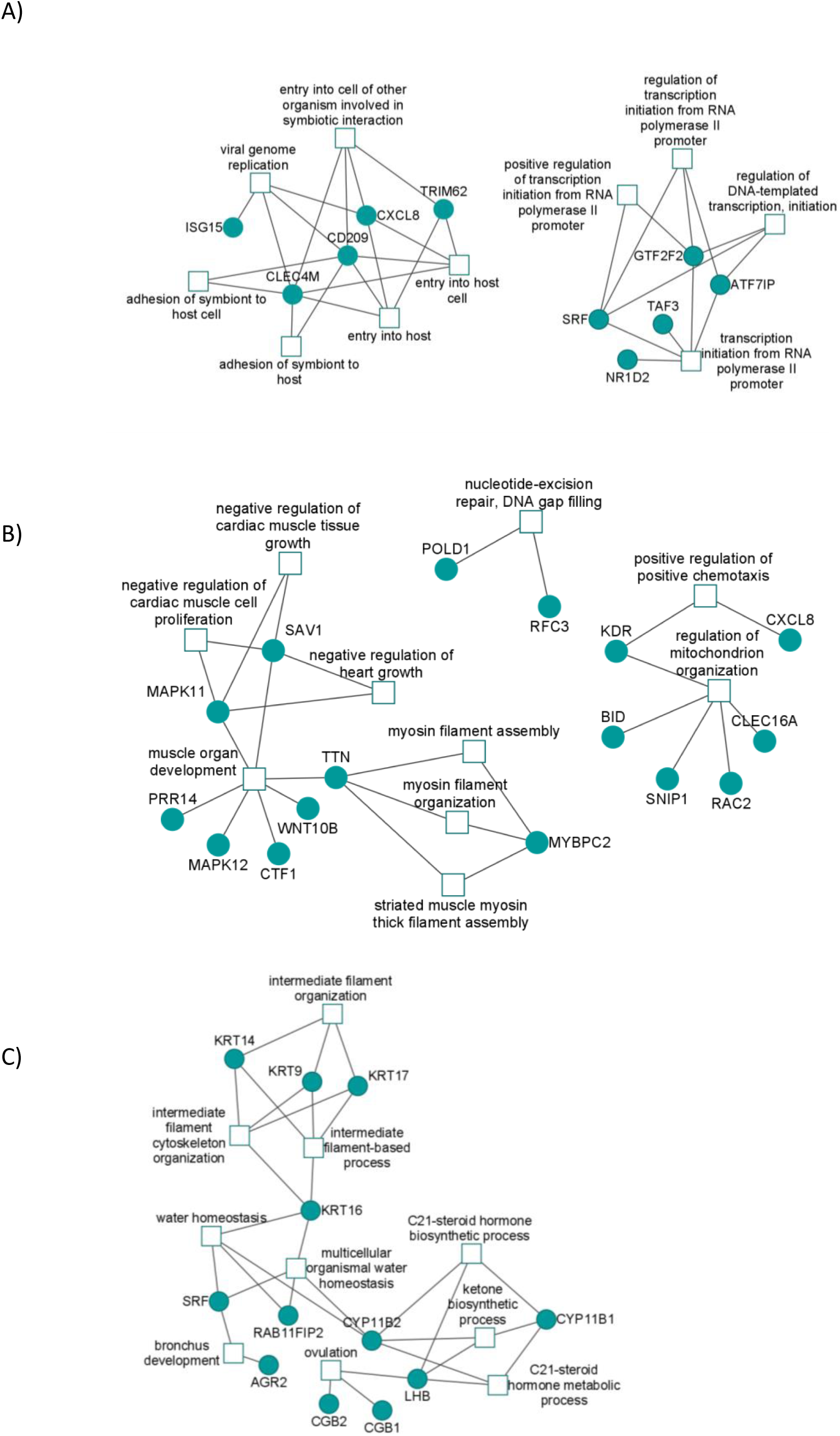
Top ten GO-terms enriched in the 100 most significant genes in gene-based analysis for A) MG, B) EOMG and C) LOMG. GO-term enrichment tests were conducted using ClusterProfiler.

### Onset-specific Genomewide Association analyses

To investigate the risk factors for each specific MG subgroup we performed onset-specific GWAS meta-analyses. Results for the individual datasets are shown in Supplement (Figures S2, S9 and S10). By setting the onset age threshold at 50 years, 455 cases and 3,508 controls were included in the EOMG GWAS, while 946 cases and 3,508 controls were analyzed in the LOMG GWAS. Genomic control factor for both groups did not indicate residual stratification (EOMG λ_GC_=1·008; LOMG λ_GC_= 1·041). SNP-heritability for EOMG was h^2^_SNP_= 0·64 (SE=0·12, P=7·81×10^−11^) and h^2^_SNP_= 0·53 (SE=0·07, P=5·55×10^−17^) for LOMG. LD-independent regions with p<10^−5^ for index SNPs (including variants within 3-Mb windows and r^2^>0·1 with index SNPs) are shown in the Supplement (Tables S3 and S4).

In EOMG GWAS, we detected 20 LD-independent genomewide significant regions (p_index-SNP_< 5×10^−8^) in the HLA region. This high number could be due to the complicated LD pattern. SNP rs9262202 was the top variant (p=5·56×10^−29^, OR=0·37, 95% CI 0·32-1·37) (Table 3, Figure 1B). It is located on chromosome 6 in the intergenic region between *DDR1*(−90·95kb) and *IER3*(+48·58kb). In the gene-based analysis excluding the HLA region - see methods), three loci, namely *SRCAP, LOC730183*, and *FBRS*, were significantly associated after correcting for 17,993 gene tests (p=2·78×10^−6^) (Figure S4B). Conditional analysis (Figure S11), showed that the only independently associated SNP (rs8058928) was located in the intergenic region of *SRCAP(−5*·22kb), *LOC730183(−4*·72kb), and *FBRS(22*·18kb).

**Table 3:**
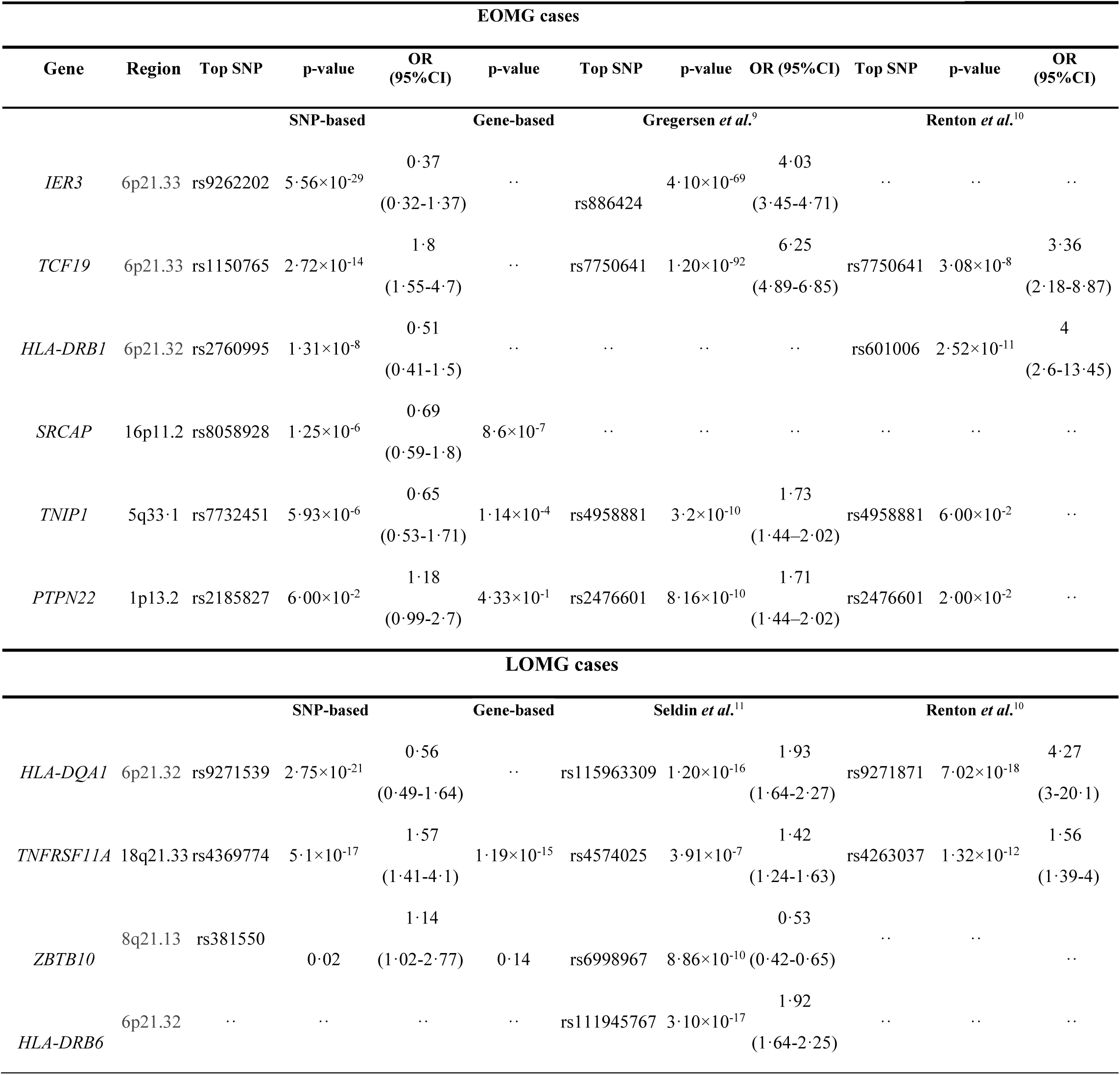
Genome-wide significant loci in EOMG and LOMG. Genetic loci detected as significantly associated with EOMG or LOMG in our study in comparison to previous results. For the non-HLA loci, SNP column corresponds to the most significant SNP nearest to each gene within 10-kb. The significance threshold for gene-based analysis is p=2·78×10^−6^. For the HLA region (6p22.1-21.3), we report the top SNP in each study and the closest gene to the SNP.

In the LOMG GWAS, five LD-independent genomewide significant regions were detected. SNP rs9271539 was the top variant (p=2·75×10^−21^, OR=0·37, 95% CI 0·49-1·64) (Table 3, Figure 1C). It is located in the intergenic region between *HLA-DQA1*(−15·15kb) and *HLA-DRB1*(+32·42kb). LOMG gene-based association analysis (excluding the HLA region - see methods), revealed *TNFRSF11A* as the only genomewide significant locus after correcting for 17,995 gene tests (p=2·78×10^−6^) (Figure S4C).

Focusing on the top 100 and 200 genes in each of our EOMG and LOMG gene-based analysis, we created GO-term clusters and associated gene networks (Figure 2B-C and S6B-C). Although not statistically significant, processes related to muscle growth and development are highlighted in EOMG analysis while steroid hormone synthesis processes are highlighted in LOMG. We did not detect any statistically significant tissue-specific enrichment of gene expression under Bonferroni correction (30 tissues: p<1·67×10^−3^; 54 tissues: p<9·26×10^−4^) (Table 2 and Figures S7B-C and 8B-C). In EOMG, sigmoid colon, brain hypothalamus and esophagus muscularis were the top tissues enriched in expression of EOMG-associated genes, whereas thyroid, brain cerebellum and pituitary were the top three tissues for LOMG (Table 2).

### HLA associations

The HLA locus was highlighted as the top locus underlying MG susceptibility. In the MG GWAS meta-analysis, rs34481484 (*HLA-DQA1)* emerged as the top and single genomewide significant locus within HLA (Table S2). In LOMG analysis, rs9271539, residing between *HLA-DQA1* and *HLA-DRB1*, was the top variant in HLA (with three additional LD-independent genomewide significant loci in the region - Table S4). In the EOMG GWAS, rs9262202, located between *DDR1* and *IER3* and 475.6kb from the nearest HLA (*HLA-C*) gene, was the top risk locus, with additional 19 LD-independent loci in HLA region arising as genomewide significant (Table S3). Given the complexity of the LD patterns at the HLA locus, we proceeded with fine-mapping efforts. Classical HLA, their amino acid sequences and SNPs were imputed for further examination. The comparison of classic HLA alleles associations between our study and previous GWAS analyses is shown in the Supplement (Table S5).

With all MG individuals analyzed jointly, we identified two associated loci, *HLA-DRB1* and *HLA-B*. Within the *HLA-DRB1* region, *HLA-DRB1*13:01* was the most strongly associated allele (p=2·6×10^−9^, OR=0·47, 95% CI 0·37-1·44). *HLA-DRB1* SerValLeu11 almost reached genomewide significance (p=5·32×10^−8^, OR=1·3, 95% CI 1·18-3·27). After conditioning for *HLA-DRB1*13:01*, the effect of *HLA-DRB1* SerValLeu11 was reduced (conditional p=7·49×10^−6^) (Figure S8). In the *HLA-B* region, the amino acid polymorphism at position 74 (p=4·87×10^−9^, OR=1·32, 95% CI 1·2-3·33) accounted for most of the SNP effect (rs1131215, p = 4·87×10^−9^, OR = 1·32, 95% CI 1·2-3·33). None of the classical HLA alleles were detected to be significantly associated. Conditioning on *HLA-B* aa74 eliminated the effect of all variants in the region (Figure S12).

In the case of EOMG, *HLA-B* was found to be the only independent locus. Conditional analyses showed that *HLA-B* Asp9 (p=2·69×10^−37^, OR=3·24, 95% CI 2·7-14·89) accounted for both the SNP effect (rs2596492: p=2·69×10^−37^, OR=3·24, 95% CI 2·7-14·89) and the classical allele effect (*HLA-B*08:01*: p=2·69×10^−37^, OR=3·24, 95% CI 2·7-14·89; *HLA-B*08*: p=2·69×10^−37^, OR=3·24, 95% CI 2·7-14·89) in the region. After conditioning for *HLA-B* Asp9 the effect of all variants was eliminated (Figure S13).

Focusing on LOMG, *HLA-DRB1* was detected as the only independent locus. The amino acids *HLA-DRB1* Val16 and Arg25 showed the same effect (p=2·48×10^−21^, OR= 0·57, 95% CI 0·51-1·66) and seem to account for most of SNPs effect (top variant rs9256943: p=1·01×10^−20^, OR=0·58, 95% CI 0·52-1·68) as well as the classical allele effect (*HLA-DRB1*03:01*: p=5·73×10^−13^, OR=0·47, 95% CI 0·39-1·47). After introducing each of the amino acid alleles separately in the regression model, the effect of the rest of the markers in *HLA-DRB1* was reduced, with *HLA-DRB1* Lys71 (p=1·72×10^−11^, OR=0·62, 95% CI 0·54-1·71, P_conditional_=1·94×10^−7^) showing the strongest association. When *HLA-DRB1* Val16 and Arg25 alleles were introduced together in the conditional analysis, the effect of all variants in the region was eliminated (Figure S14).

### Genetic correlation with autoimmune diseases

We analyzed publicly available European-ancestry GWAS summary statistics datasets for 12 autoimmune disorders to investigate genetic correlation to MG. To ensure interpretability and power of our analysis, we included only datasets with a heritability z-score >4. By setting the p-value significance threshold at 0·05, we detected strong statistically significant genetic correlation between MG and Type 1 Diabetes (rg=0·67, SE=0·13, p=4·78×10^−7^), Rheumatoid Arthritis (rg=0·5, SE=0·12, p=3·83×10^−5^) and late-onset Vitiligo (rg=0·33, SE=0·15, p=0·03) (Figure 3; Table S1).

**Figure 3:**
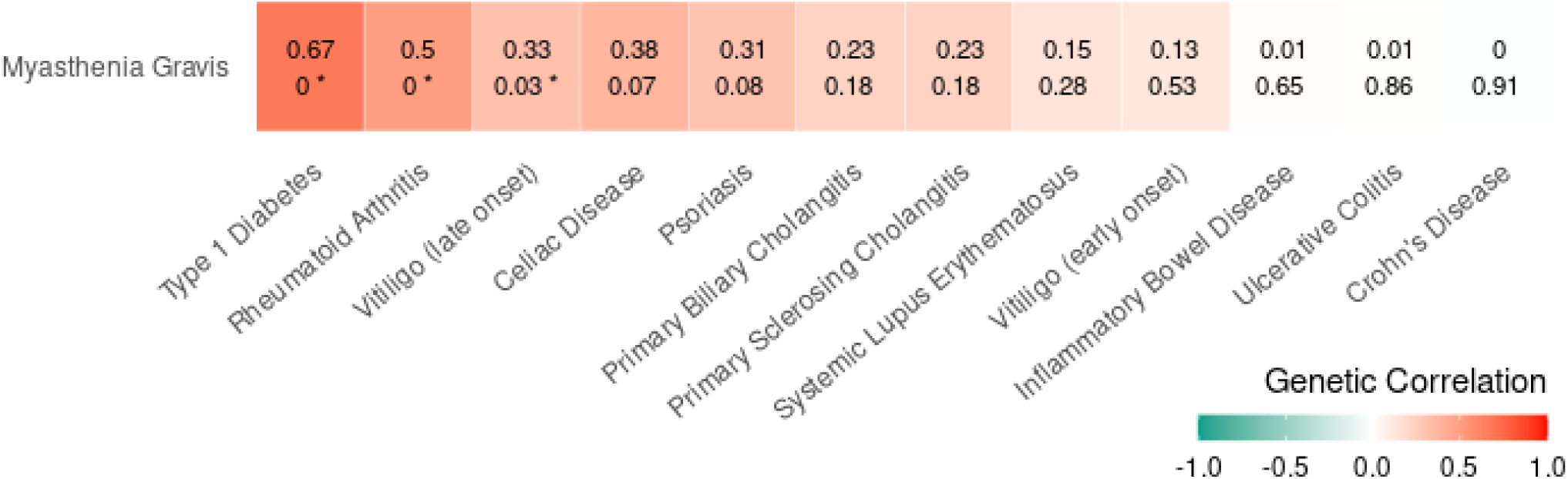
Genetic correlation of MG GWAS with GWAS for other autoimmune disorders. Analysis was performed using LDSC. Only studies with heritability z-score >4 were used. The common SNPs with the provided HapMap3 reference and the LD scores calculated from the European 1000Genomes samples were used. The upper value corresponds to the genetic correlation and the lower value corresponds to the p-value for the genetic correlation. Based on the correlation direction, the color of each box varies from green (negative genetic correlation) to red (positive genetic correlation). The asterisks denote p-values <0·05.

## Discussion

We present a GWAS meta-analysis of 1,401 MG cases and 3,508 neurologically healthy controls of European ancestry which represents the largest MG genomewide dataset analyzed to date. Our increased sample size allows us to identify several novel risk loci associated with MG and MG subtypes and replicate association with previously associated loci. We also provide clues to the pathways that underlie MG susceptibility and shed light into the genetic architecture of EOMG in comparison to LOMG.

Among the novel MG risk loci, association to *AGRN* is particularly intriguing. *AGRN* encodes the ∼200-kD proteoglycan agrin involved in the activation and synaptic stabilization of the NMJ by mediating AChR clustering. Agrin is released in the synaptic cleft from motor neuron terminals, interacts with its receptors, α-dystroglycan^27^ and LRP4,^3^ and activates the MuSK receptor. Consequently, MuSK becomes activated via autophosphorylation, thus inducing a signalling cascade which results in the clustering of various molecules involved in signal transmission at the NMJ. Clustering of AChRs occurs with the help of rapsyn,^28^ whereas synaptic stabilization occurs through agrin-mediated α-dystroglycan interaction with cytoskeleton proteins.^29^ Agrin-deficient knockout mice have normal AChR levels, but fewer postsynaptic clusters of reduced size and density,^28^ whereas immunization of mice with agrin induced MG-like symptoms.^30^ Anti-agrin autoantibodies have been detected mostly in seropositive MG patients,^31^ whereas their absence from healthy controls or patients with other neurological diseases, supports their diagnostic value as MG-specific autoantibodies.^4^ Mutations in *AGRN* are a rare cause of congenital myasthenic syndrome characterized by fatigable weakness of skeletal muscle,^32^ highlighting that agrin is a critical organizer of postsynaptic differentiation.

As also suggested by previous GWAS in EOMG and LOMG, we find different genetic architecture in each subgroup ^10,11^. *TNFSRF11A* variants emerge at the top among our hits, demonstrating an even stronger association signal in LOMG. On the contrary, this gene was not found significantly associated with EOMG. Similarly, *CTLA4* was previously associated with the general MG phenotype and LOMG,^33,34^ both in candidate gene association studies and at the genomewide level.^10^ We too, report a significant association of *CTLA4* to MG in our gene-based meta-analysis (p_adj_=1·43×10^−6^), but this association did not pass significance level for LOMG (p_adj_=3·65×10^−6^). Previously, the *CTLA4* association was not replicated in a GWAS of Asian MG cases,^35^ which could be explained by the different ethnic background or the small sample size (109 MG cases), nor in a European LOMG GWAS (532 LOMG cases).^11^ In our EOMG GWAS, the previously reported association to *TNIP1*^*9*^ only reached a suggestive p value. Furthermore, we did not find evidence supporting the previously reported implication of *PTPN22* in EOMG and *ZBTB10* in LOMG.^9,11^ Importantly, we detected a novel genomewide significant association with EOMG at a region that lies between the *SRCAP* and *FBRS* genes.

In our study, HLA fine-mapping revealed two independent regions (*HLA-DRB1* and *HLA-B)*, contributing to MG susceptibility. The *HLA-DRB1*13:01* allele was the source of the association signal in *HLA-DRB1* and its protective effect has been previously reported in both EOMG and LOMG^7,36^ as well as in Rheumatoid Arthritis (RA).^37^ In *HLA-B*, an amino acid residue at position 74 was found to drive the signal conferring risk to MG. Additional onset-specific analyses revealed the classical alleles *HLA-B*08:01* and *HLA-B*08* to be predisposing in EOMG, in consistency with previous studies^7–9^ (Table S5). We identified that aspartate at position 9 in *HLA-B*, which resides within the peptide-binding groove, correlates with the association of *HLA-B*08*, possibly impairing peptide loading and presentation, as shown in RA, as well.^38^ In LOMG, the *HLA-DRB1*03:01* was the most strongly associated classical allele with a protective role against MG, as confirmed by previous studies^11,36^ (Table S5). At the amino acid level, lysine at position 71 exceeded the significance of all other association signals in *HLA-DRB1* and is located in the “shared epitope” (a common amino acid sequence at positions 70-74 at the extracellular domain of the DRβ1 chain), which constitutes a main risk factor for RA susceptibility.^39^

Intriguingly, GO analysis highlights the GO cluster of symbiotic processes among the top enriched in our MG hits. Symbiotic intestinal microbiota represent a central regulator of host immune homeostasis; it is now clear that immunomodulatory molecules produced by commensal bacteria drive the host’s immune system to mature.^40^ Interestingly, an altered fecal microbiota pattern has been recently found to characterize seropositive MG patients.^41^ Zheng et al. identified that a panel of microbes correlated with aspects of MG severity and that gut microbial and fecal metabolite markers enabled discrimination of MG cases from healthy controls with 100% accuracy.^42^ Administration of probiotic strains in an experimental rat model of MG ameliorated clinical and laboratory measurements.^43^ Moreover, colonization of germ-free mice with human MG microbiota resulted in impaired locomotion ability and an increased inflammatory cytokines profile compared to colonization with healthy human microbiota,^42^ suggesting that gut microbiome may contribute to the development of MG, presumably by modulating host metabolism. ^42^ Colon sigmoid was the most over-represented tissue in EOMG GWAS, however it did not reach statistical significance. Such preliminary evidence suggests a role for gut microbiome in MG susceptibility warranting further studies.

Motivated by the clinical observation of frequent comorbidity of other autoimmune conditions in MG patients,^44^ we sought to investigate the potential genetic correlation of MG GWAS to GWAS for other autoimmune disorders. About 15% of MG patients have been reported to have a second autoimmune condition.^44^ A high genetic correlation would suggest the possibility of shared pathways of pathogenesis.^45^ We detected significant genetic correlation between MG, Type 1 Diabetes, Rheumatoid Arthritis and late-onset Vitiligo. To our knowledge, MG has not yet been included in autoimmune cross-disorder meta-analyses. Our results show that such a cross-disorder genetic analysis is warranted and could potentially shed light on a shared genetic mechanism between MG and other autoimmune disorders.

In conclusion, the present meta-analysis provides support for the involvement of *AGRN* in MG etiology and strong support for the involvement of previously identified loci including HLA, *TNFSRF11A* and *CTLA4*. Furthermore, our findings underline the fact that a different genetic architecture may distinguish EOMG from LOMG with processes related to muscle development and growth highlighted in EOMG and steroid hormone biosynthesis pathways in LOMG. Perhaps variations at specific genetic loci, create a milieu predisposing to the disease, upon which different environmental, hormonal and sex-specific factors determine each subgroup’s susceptibility to disease onset and severity. Although, this study represents the largest MG GWAS meta-analysis to date, still larger sample sizes of individual subgroups will be required in order for genetics to provide a robust explanation for their distinct immunological, histological and epidemiological characteristics.

## Data Availability

Summary statistics will become available upon publication of this study at the Paschou lab website. De-identified raw genotype data can become available upon request via email to the corresponding author.

## Acknowledgements

All study participants are warmly thanked for consenting to participate in the study. Dr. Iordanis Karagiannidis is acknowledged for his dedicated work with the curation of the Greek cohort. This work was supported by NSF award #1715202, the European Social Fund and Greek funds through the National Strategic Reference Framework (NSRF) THALES Programme 2012–2015 and the NSRF ARISTEIA II Programme 2007–2013 to PP, and grants from the Association Francaise contre les Myopathies (AFM, Grant No. 80077) to ST.

## Author Contributions

PP and MG conceived and designed the study, PP, MG, AT, ZZ, FT acquired samples and data, PP, AT, AF, MM, FT, ZY analyzed data, PP, MG, ZZ, FT, YC, GL, EY, EP, KP, JT, KK, EM, SP, NP, DP, PP, AR, MT, ET, XT, JY, JS, KK, ST contributed data acquisition, sample collection and clinical characterization, all authors contributed to drafting the manuscript, providing critical comments and revisions.

