## Supplementary Text and Figures for "A Myasthenia Gravis genomewide association study of three cohorts identifies Agrin as a novel risk locus"

### Supplementary Methods

#### Samples

Following quality control, we integrated three different GWAS datasets for a total of 1,401 cases and 3,508 controls: Our data included 1) 196 Greek and Greek-Cypriot MG cases and 1,057 ancestry-matched controls, 2) 964 European-American MG cases and 1,985 ancestry-matched controls, available through dbGaP, and 3) 241 MG cases and 466 controls, available through the UK biobank.

**Greek-Cypriot sample:** A total of 211 MG cases were recruited in Greece and Cyprus between 2010-2017. Study participants were temporarily diagnosed with possible MG based on clinical evidence (neurological evaluation of muscle involvement). Of those, 73 cases were characterized as early-onset (30 y.o average age at diagnosis, 23.29% males) and 138 were late-onset (67 y.o average age at diagnosis, 66,67% males). Cases were then referred to either the Hellenic Pasteur Institute or Tzartos NeuroDiagnostics or the Department of Pharmacy of the University of Patras where their status was confirmed by quantitative determination of the presence of anti-AChR autoantibodies using a commercially available radioimmunoassay (RiaRSR™ AChRAb kit, RSR Ltd., Cardiff, UK). Results were considered positive for the presence of anti-AChR antibodies if ≥0.5 nmol/L. Anti-MuSK and anti-LRP4 positive MG cases were not included in this sample. A total of 1,289 population-based controls had been previously collected from multiple sites in Greece and Cyprus, of which 51,71% were male. All participating individuals gave written informed consent. The study was approved by the Ethics Committee of the Hellenic Pasteur Institute and by the National Bioethics Committee of Cyprus (Bioethics Committee for the Evaluation of Biomedical Research in Humans - EEBK/EP/2016/20) and was in accordance with the Helsinki Declaration of Human Rights.

**dbGaP dataset:** The MG-dbGaP dataset consisted of data for 1,032 cases collected from multiple sites in North America and 1,998 controls. The recruitment criteria have been previously described.[1]

**UK Biobank dataset:** We analyzed data for a total of 241 individuals of European ancestry with the G70.0 phenotype (Myasthenia Gravis) in the ICD-10 diagnoses data-field. As controls, we initially considered the three closest neighbours for each case based on the Euclidean distance calculated from the first two PCs of the genomic dataset (765,067 SNPs as described below). Among the selected control individuals, we kept only those who did not report any long-standing illness, disability or infirmity.

**Genotyping**

**Greek-Cypriot dataset:** All of the 211 cases and 562 out of 1,289 controls were genotyped on Illumina PsychArray-24v1.1 BeadChip that covers 593,260 markers. The remaining 727 controls[2] were genotyped on Illumina Infinium Omni2.5, which targets approximately 2,381,000 markers.

**dbGaP dataset:** The European-American cases were genotyped using the Illumina HumanOmniExpress BeadChip, covering about 730,525 markers. Controls were genotyped on Illumina HumanOmni1-Quad BeadChip capturing approximately 1,140,000 markers.

**UK Biobank dataset:** The UK Biobank samples were genotyped either on Affymetrix UK BiLEVE Axiom or Affymetrix UK Biobank Axiom® array. Both platforms target about 850,000 variants and have more than 95% overlap.

#### Quality Control

To ensure high quality of the analyzed datasets, we applied stringent quality control criteria. Using PLINK 1.9[3], poorly genotyped individuals (missingness >2%), individuals with high heterozygosity deviations and those with sex discrepancies were removed. Identity-by-Descent (IBD) analysis was performed in order to detect cryptic relatedness between samples and one individual of each pair with pi-hat values above 0·1875 was removed. Principal Component Analysis (PCA) using EIGENSOFT SMARTPCA[4] was conducted to remove individuals that were considered as outliers based on ancestry.

Before proceeding to genotype imputation, each genotyped batch was aligned on HRC reference using the toolbox provided at http://www.well.ox.ac.uk/~wrayner/tools/. For the Greek and Cypriot controls an additional round of batch effect testing was performed. Consequently, the different genotyped batches of each dataset were merged using only shared SNPs between them. The final dataset consisted of 245,075 SNPs for the Greek-Cypriot sample, 626,761 SNPs for the European-American sample and 721,950 SNPs for the UK Biobank sample.

#### Imputation and genetic association analyses

The Michigan Imputation Server pipeline[5] was used to perform the genotype imputation with HRC r1.1 as the reference panel. In each dataset, SNPs with imputation accuracy $r^2$ <0·7, genotype posterior probability GP <0·8 and minor allele frequency (MAF) <0·01 were excluded.

Genome-wide association tests were performed for each dataset in PLINK 1.9 using the logistic regression under an additive model including the principal components (PCs) suggested by SMARTPCA as covariates: PC1 for the Greek-Cypriot dataset and PC7 for the European-American dataset, while no significant PCs were identified for the UK Biobank dataset owing to our careful matching of cases and controls during sample selection. The GWAS meta-analysis was conducted on summary statistics of each dataset using the inverse-variance method in METAL[6]. Heterogeneity was assessed with Cochran's $I^2$ statistic and only SNPs present in all three studies with MAF >0·01 in each study and heterogeneity p-value >0·05 were included in downstream analyses. Linkage disequilibrium (LD) score regression (LDSC)[7] analysis was performed on SNPs with INFO score >0·9 and MAF >0·01 in each dataset, to examine whether there are residual artifacts or stratification.

#### Conditional analyses

Conditional analyses were performed with GCTA-COJO[8] to identify the independently associated SNPs. As a reference sample, we used the shared set of the three datasets; a stepwise selection model with a p-value threshold of $10^{-5}$ was selected to identify the independent loci.

#### GO terms enrichment

We used ClusterProfiler[9] to perform enrichment tests for Biological Process GO clusters using the top 100 genes from the gene-based analysis. We used org.Hs.eg.db to map the genes to the GO-terms and evaluated the enrichment through a hypergeometric test.

### Supplementary Figures

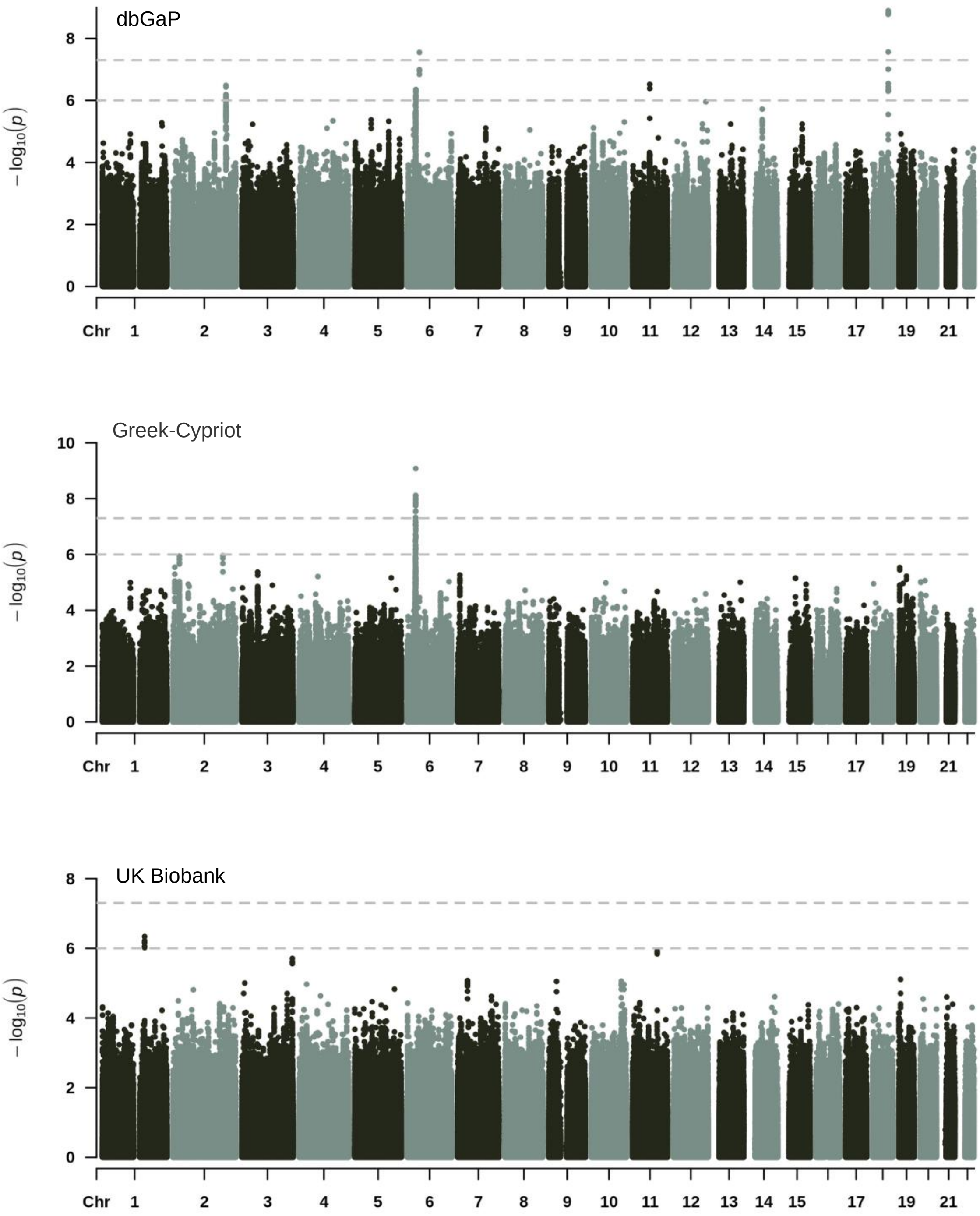

**Supplementary Figure 1: GWAS in individual MG datasets.** The dbGaP dataset included 964 MG cases and 1,985 controls; PC 7 was used in the association model as covariate. The Greek-Cypriot post-QC dataset included 196 MG cases and 1,057 controls; PC 1 was used in the association model as covariate. The UK Biobank dataset included 244 MG cases and 466 controls; no correction for PCs was needed thanks to careful pre-selection of controls to match the ancestry of the cases.

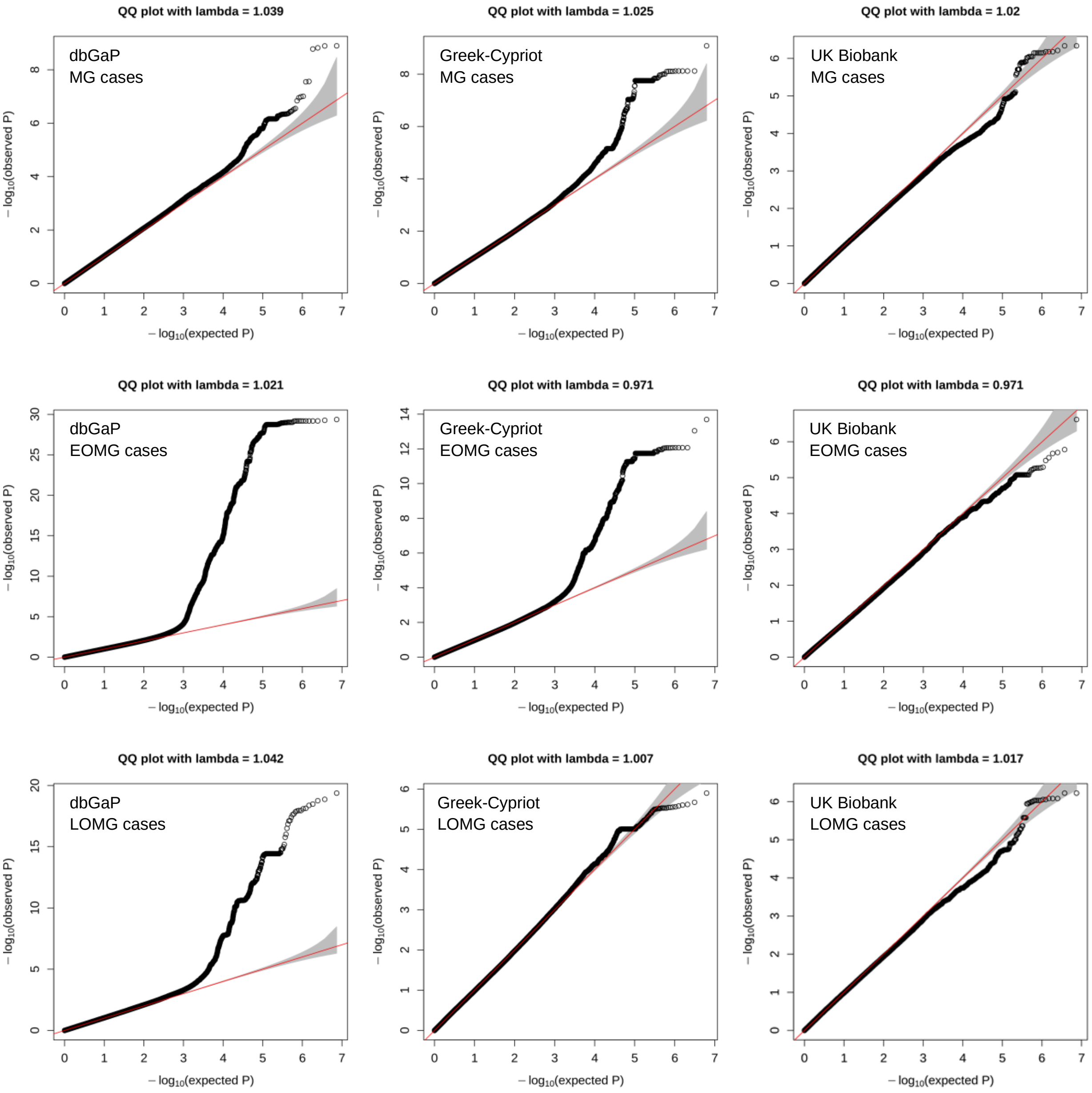

**Supplementary Figure 2: Quartile-quartile plots of the distribution of expected vs observed P values of the GWAS analyses of the individual datasets.** The 95% confidence interval of expected values is indicated in gray.

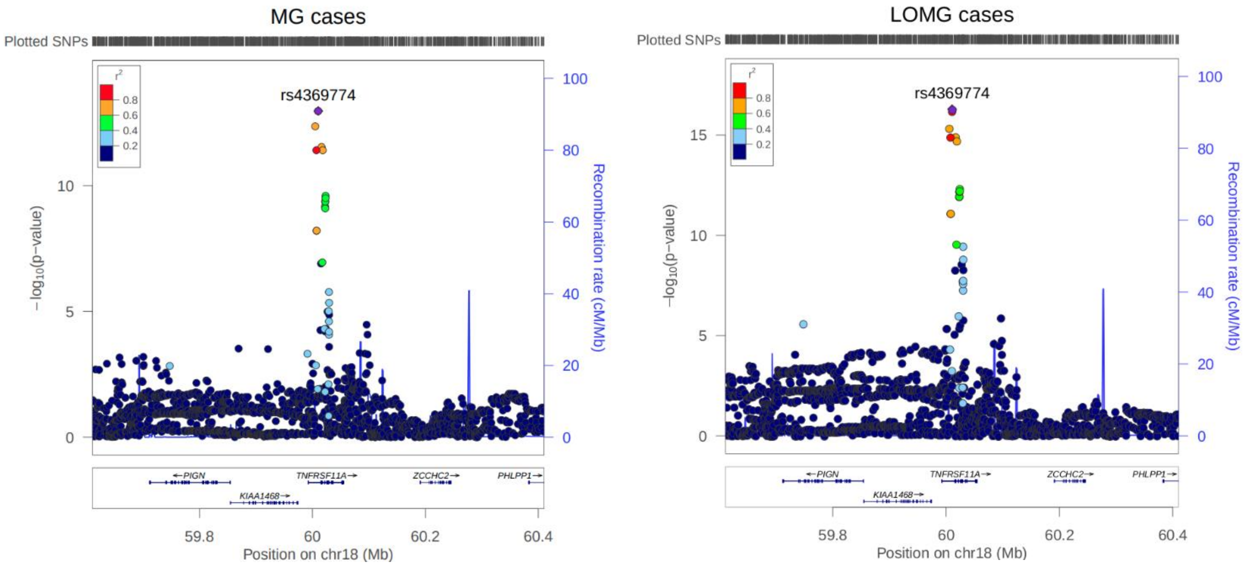

**Supplementary Figure 3: Regional association plot of the signal on chromosome 18 for MG cases and LOMG cases separately.** The different colors indicate the $r^2$ of each variant with the top SNP rs4369774. The plots were generated using LocusZoom.

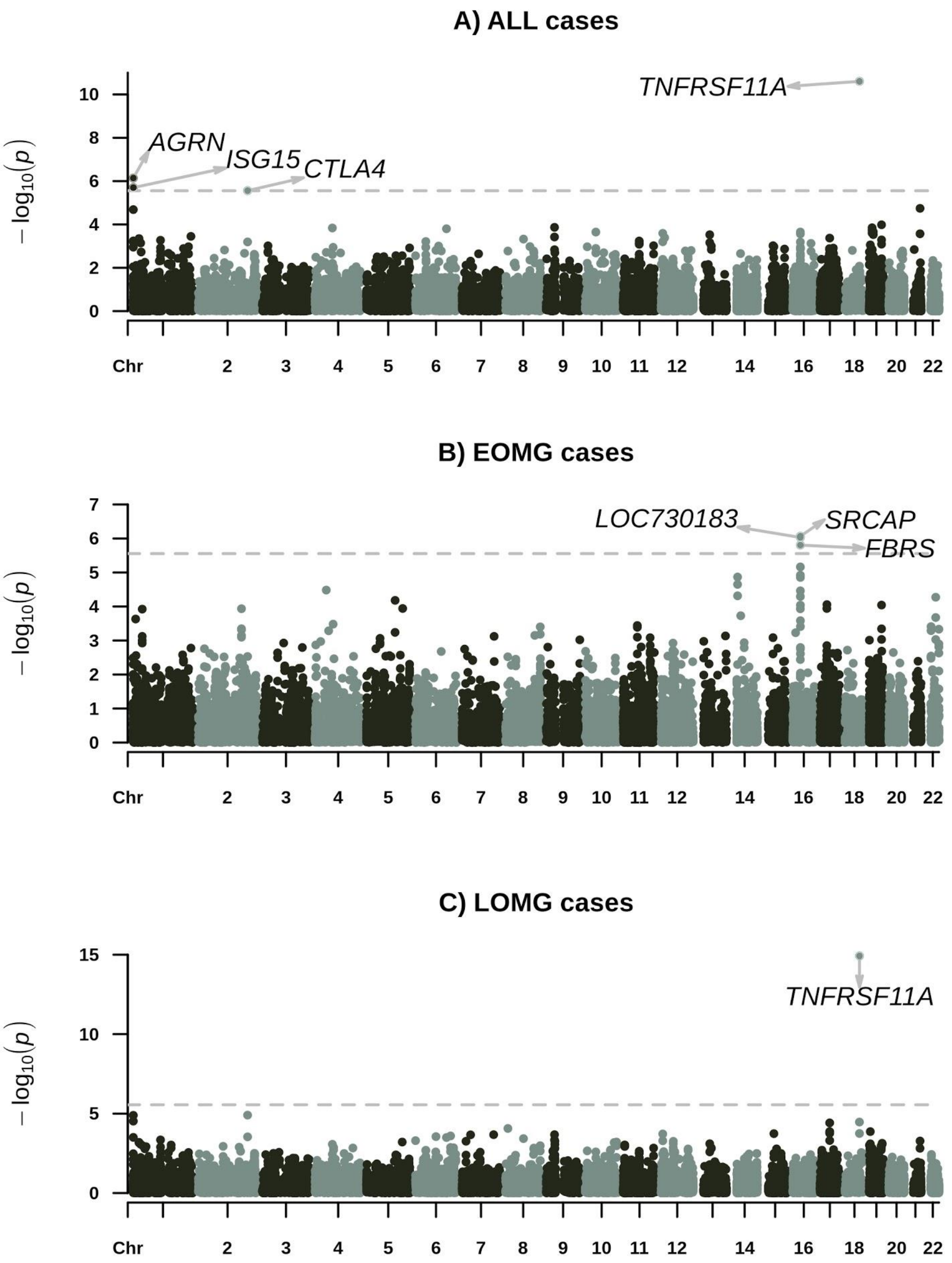

**Supplementary Figure 4: Gene-based analyses for MG GWAS (excluding the HLA locus).** The analysis was run as implemented in MAGMA with a 20kb flanking window around the genes in MAGMA. The extended HLA region (24-33Mb) on chromosome 6 was excluded from analyses. A) Gene-based analysis for 1,401 MG cases and 3,508 controls. The dashed line indicates the threshold of significance ($p=2{\cdot}78\times10^{-6}$) after Bonferroni correction for 17,994 tests. B) Gene-based analysis for 455 EOMG cases and 3,508 controls. The dashed line indicates the threshold of significance ($p=2{\cdot}78\times10^{-6}$) after Bonferroni correction for 17,993 tests. C) Gene-based analysis for 946 LOMG cases and 3,508 controls. The dashed line indicates the threshold of significance ($p=2{\cdot}78\times10^{-6}$) after Bonferroni correction for 17,995 tests.

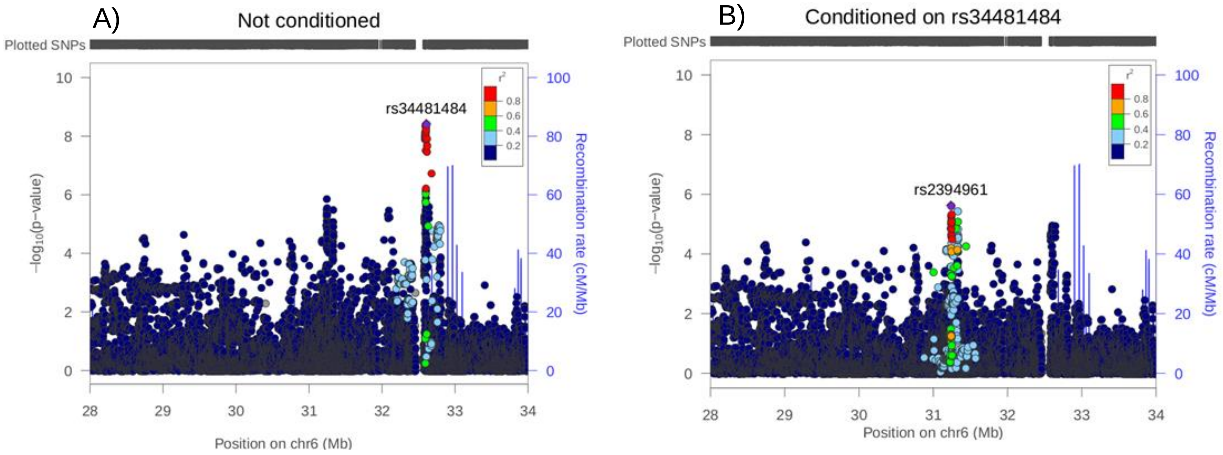

**Supplementary Figure 5: Regional association plot of the signal on chromosome 1. A)** Without conditioning, **B)** conditioning on rs3128125. The conditional analyses were performed using GCTA-COJO to identify the independently associated SNPs; the p-value threshold of $10^{-5}$ was selected to identify the independent loci. GCTA-COJO identified rs3128125 as the only independent signal. The plots were generated using LocusZoom.

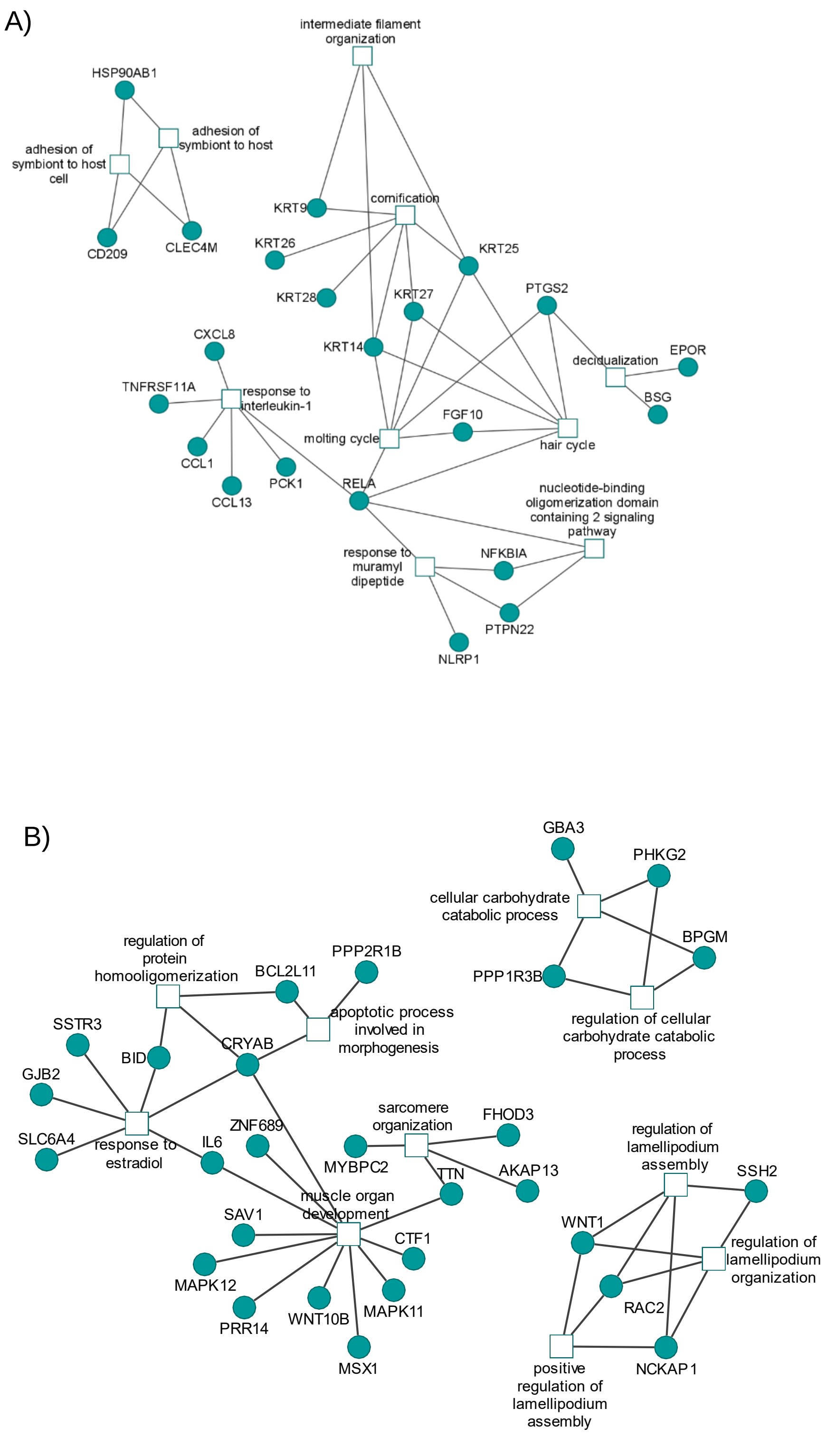
A)
intermediate filament organization
HSP90AB1
adhesion of symbiont to host
adhesion of symbiont to host cell
CD209
CLEC4M
KRT9
cornification
KRT26
KRT25
KRT28
KRT27
PTGS2
KRT14
CXCL8
TNFRSF11A
response to interleukin-1
decidualization
EPOR
BSG
FGF10
molting cycle
hair cycle
CCL1
CCL13
PCK1
RELA
nucleotide-binding oligomerization domain containing 2 signaling pathway
response to muramyl dipeptide
NFKBIA
PTPN22
NLRP1
B)
GBA3
PHKG2
cellular carbohydrate catabolic process
BPGM
PPP1R3B
regulation of cellular carbohydrate catabolic process
regulation of protein homooligomerization
BCL2L11
PPP2R1B
apoptotic process involved in morphogenesis
SSTR3
BID
CRYAB
GJB2
SLC6A4
response to estradiol
IL6
ZNF689
sarcomere organization
FHOD3
MYBPC2
AKAP13
TTN
muscle organ development
SAV1
CTF1
MAPK12
PRR14
WNT10B
MAPK11
MSX1
regulation of lamellipodium assembly
SSH2
WNT1
regulation of lamellipodium organization
RAC2
positive regulation of lamellipodium assembly
NCKAP1

C)

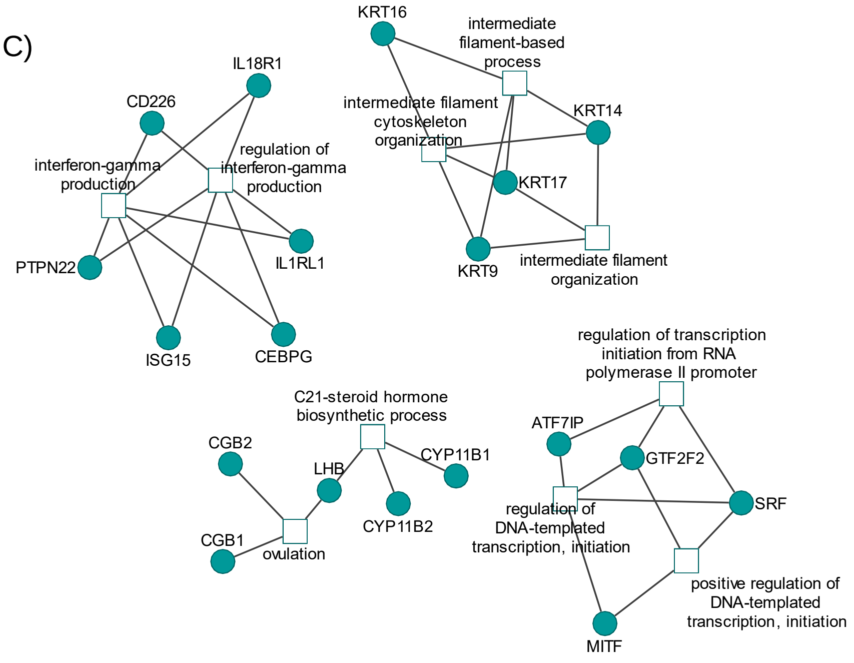

**Supplementary Figure 6:** Network of the top ten enriched GO-terms based on the 200 most significant genes in the gene-based analysis (excluding HLA region) for A) MG cases, B) EOMG cases and C) LOMG cases.

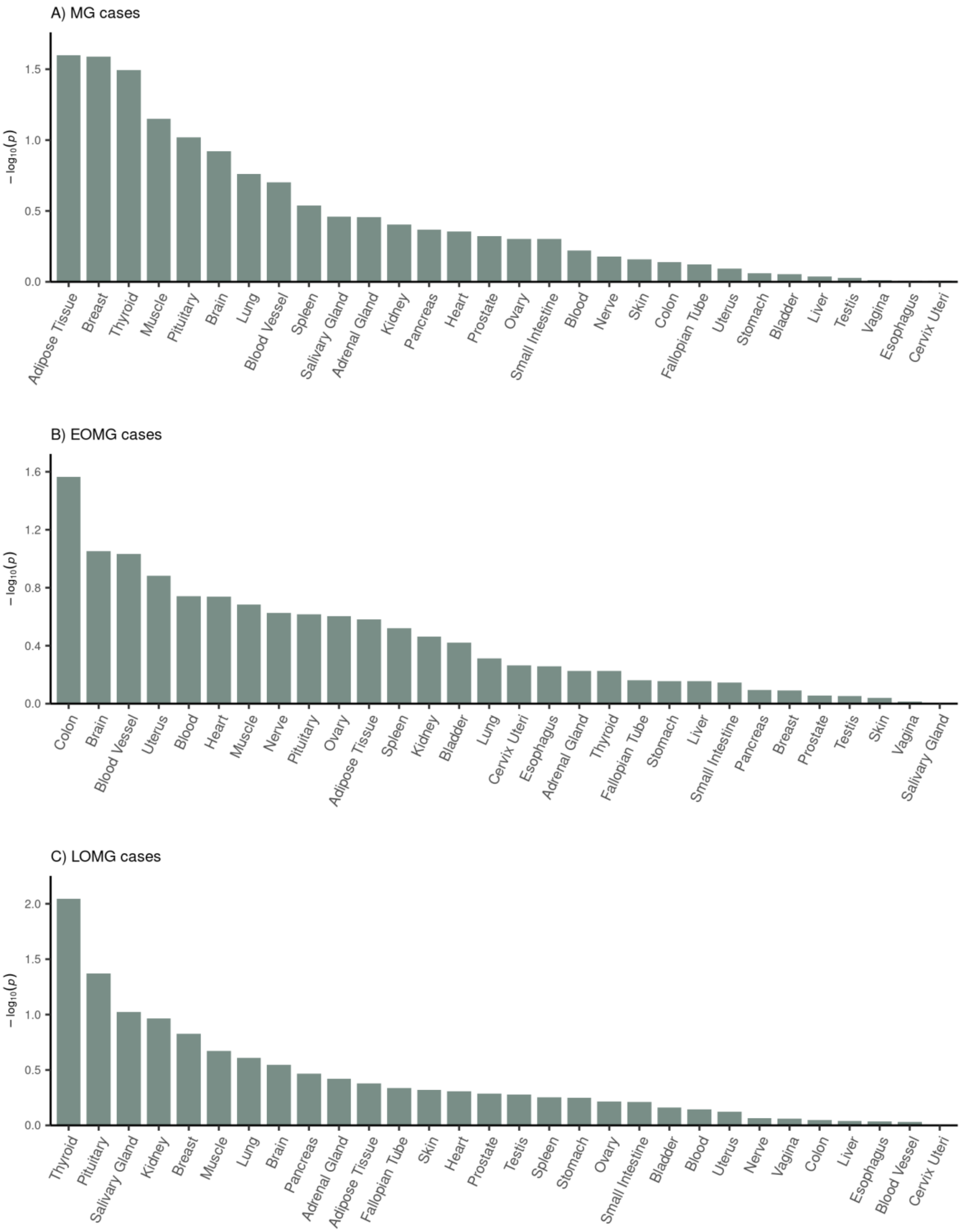

**Supplementary Figure 7: Tissue-specific enrichment analysis of genes associated with MG in 30 general human tissues.** Analysis was performed in FUMA's SNP2GENE mode; the tissue-specific enrichment was tested in 30 general human tissue types from 948 donors using GTEx v.8 RNA-seq data. The HLA region was excluded from gene-based analysis. The significance threshold for the tissue-specific test was calculated using the Bonferroni correction for 30 tests ($p=1.67 \times 10^{-3}$). Panels A, B and C, show results from the MG, EOMG and LOMG cases, respectively.

A) MG cases

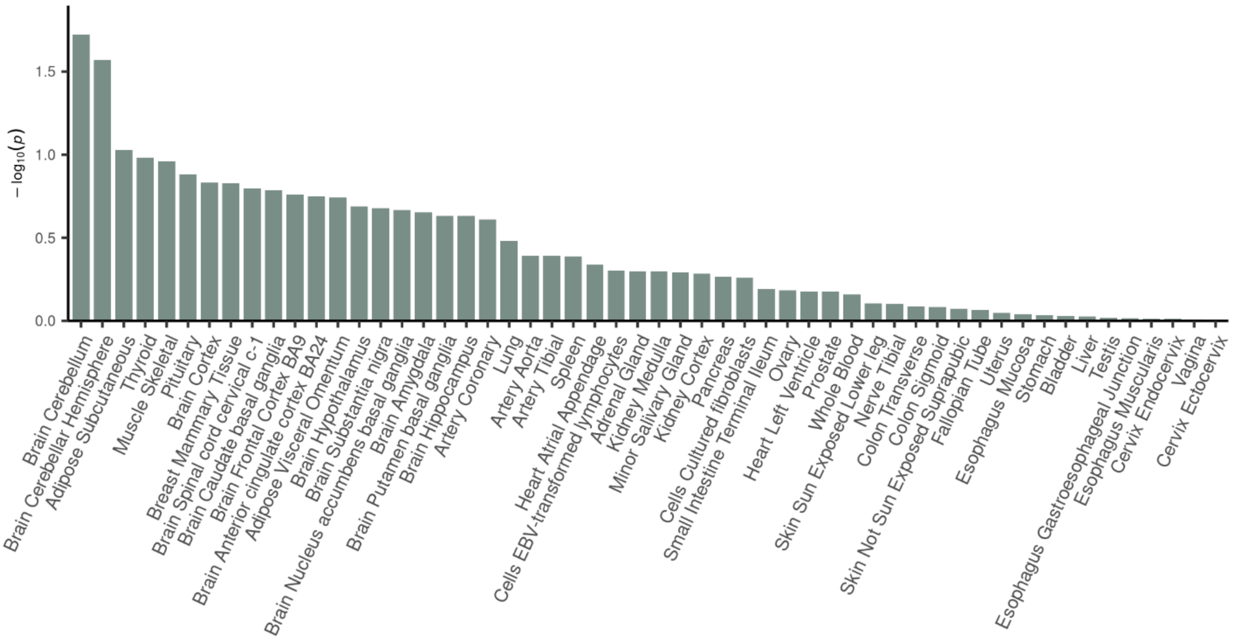

B) EOMG cases

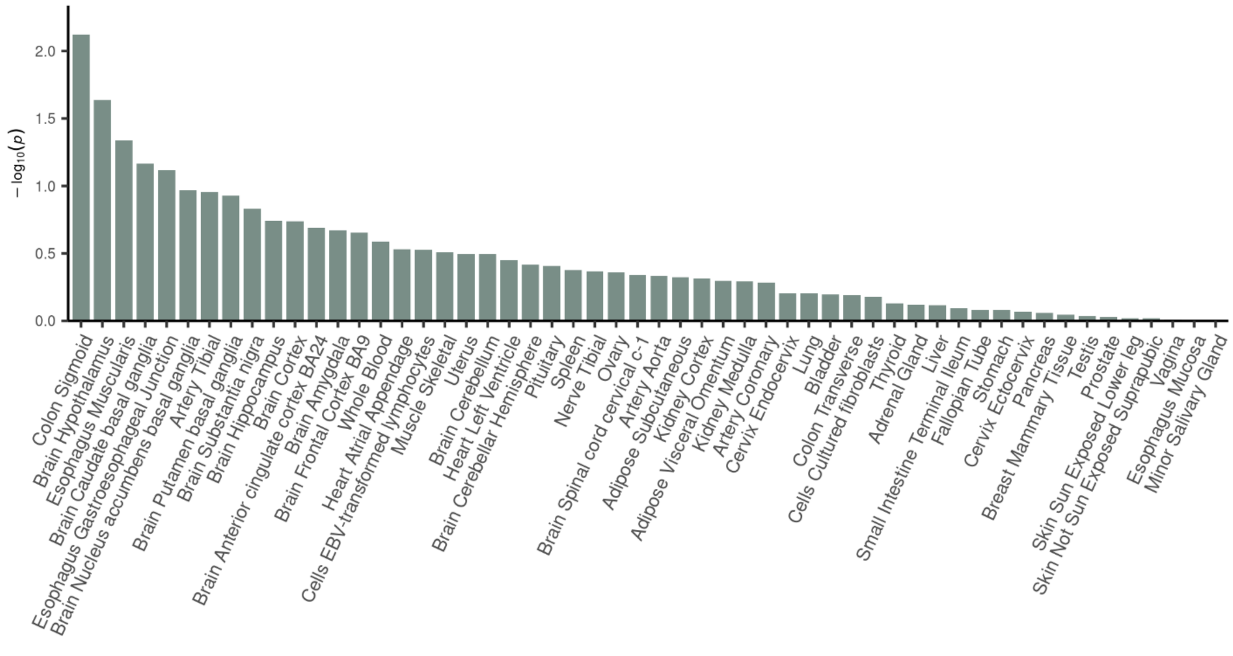

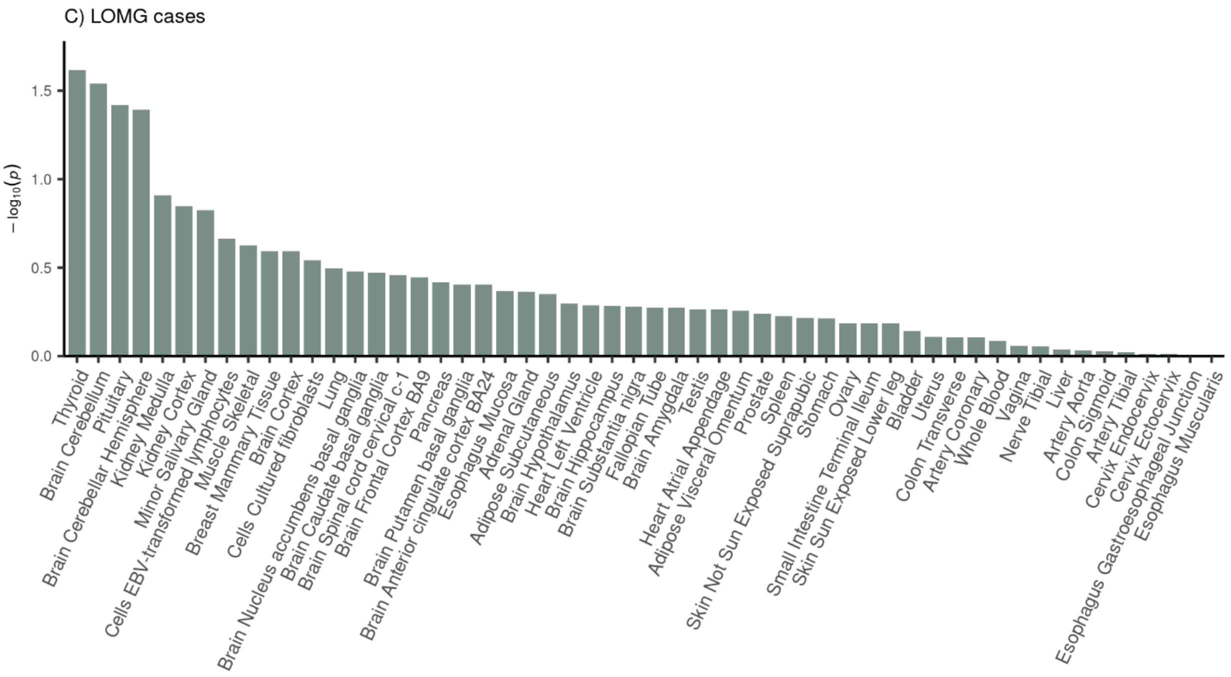

**Supplementary Figure 8: Tissue-specific enrichment analysis of genes associated with MG in 54 human tissues.** Analysis was performed in FUMA's SNP2GENE mode; the tissue-specific enrichment was tested in 54 human tissue types from 948 donors using GTEx v.8 RNA-seq data. The HLA region was excluded from gene-based analysis. The significance threshold for the tissue-specific test was calculated using the Bonferroni correction for 54 tests (p=9·26x10-4). Panels A, B and C, show results from the MG, EOMG and LOMG cases, respectively.

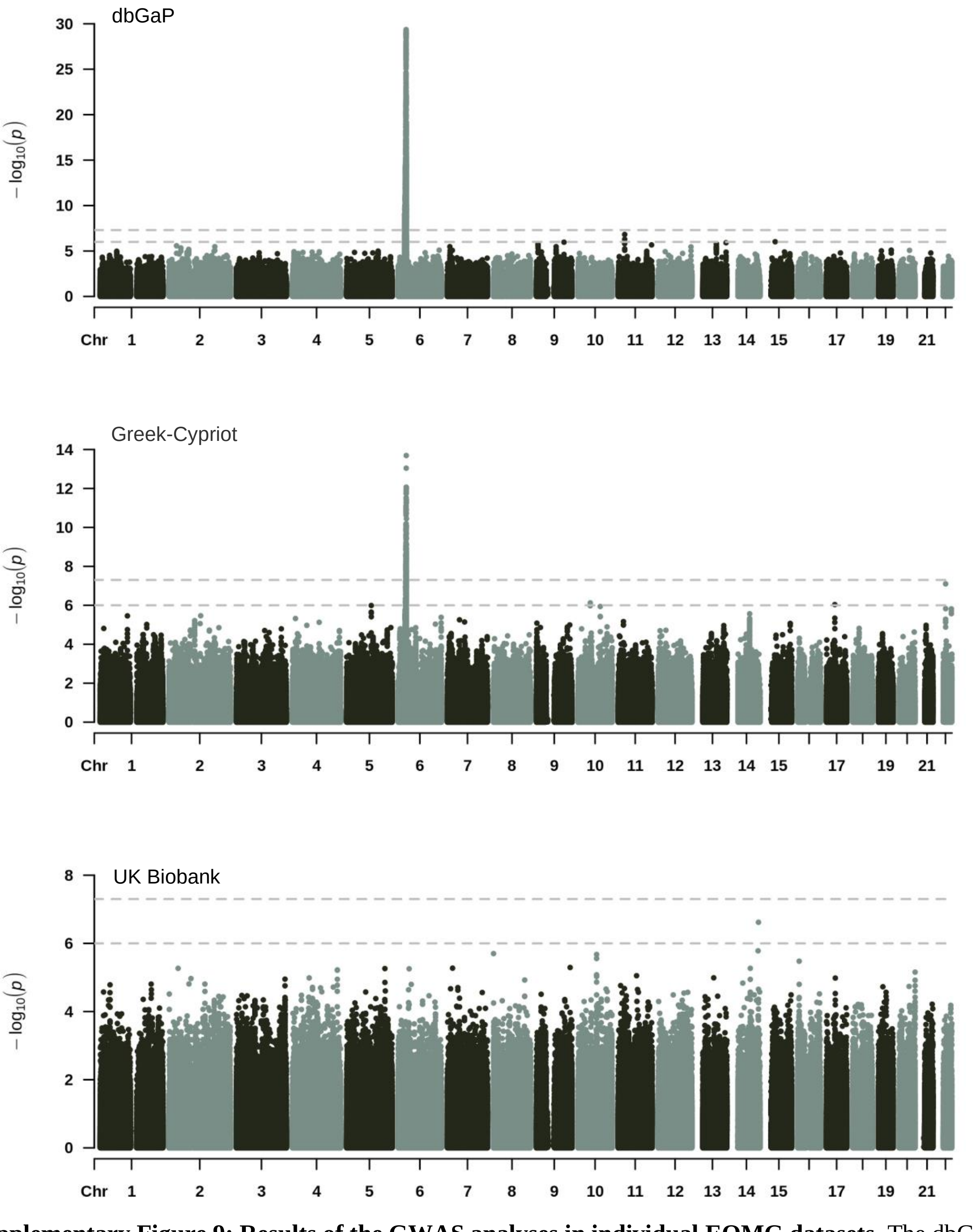

**Supplementary Figure 9: Results of the GWAS analyses in individual EOMG datasets.** The dbGaP dataset included 322 EOMG cases and 1,985 controls; PC 7 was used in the association model as covariate. The Greek-Cypriot dataset included 66 EOMG cases and 1,057 controls; there was no correction for PCs required. The UK Biobank dataset included 67 EOMG cases and 466 controls; no correction for PCs was required.

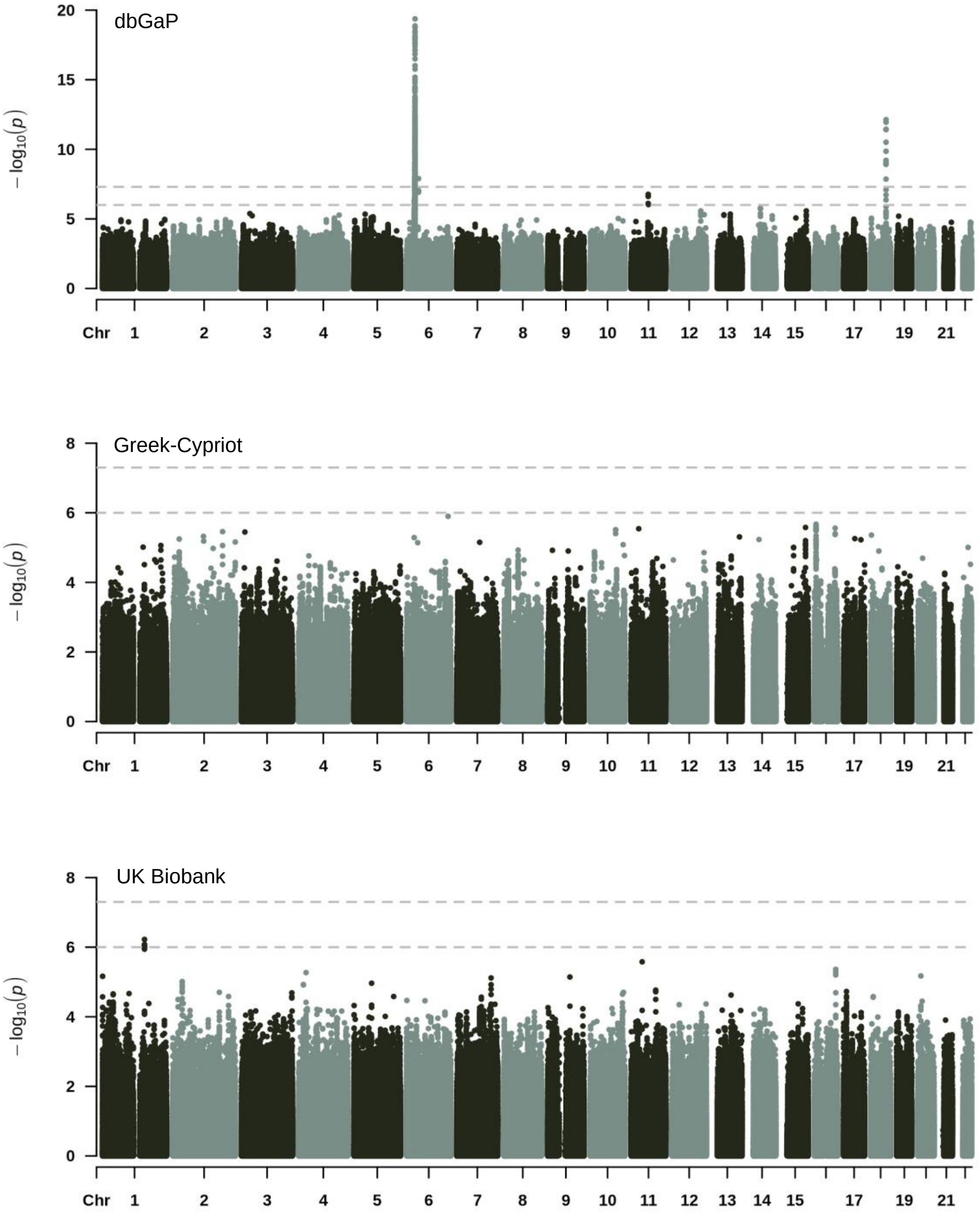

**Supplementary Figure 10: Results of the GWAS analyses in the individual datasets of LOMG cases.** The dbGaP dataset included 642 LOMG cases and 1,985 controls; PC 7 was used in the association model as covariate. The Greek-Cypriot dataset included130 LOMG cases and 1,057 controls; PC 1 was used in the association model as covariate. The UK Biobank dataset included 174 LOMG cases and 466 controls; there was no required correction for PCs.

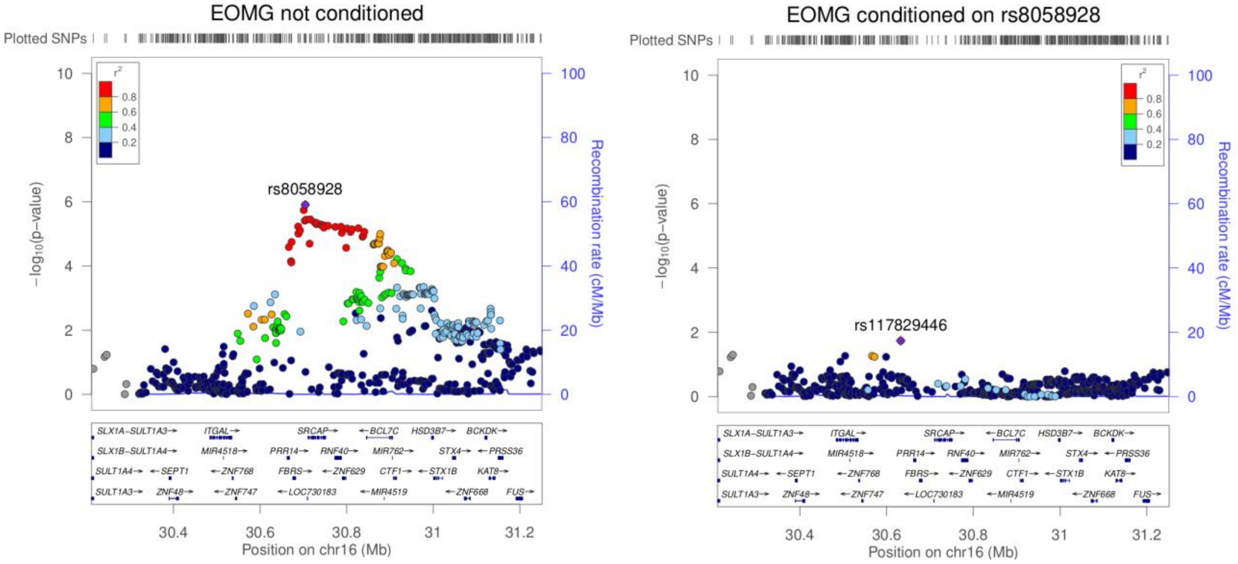

**Supplementary Figure 11: Regional association plot of the signal on chromosome 16. A)** Without conditioning, **B)** conditioning on rs8058928. The conditional analyses were performed using GCTA-COJO to identify the independently associated SNPs; the p-value threshold of $10^{-5}$ was selected to identify the independent loci. GCTA-COJO identified rs8058928 as the only independent signal. The plots were generated using LocusZoom.

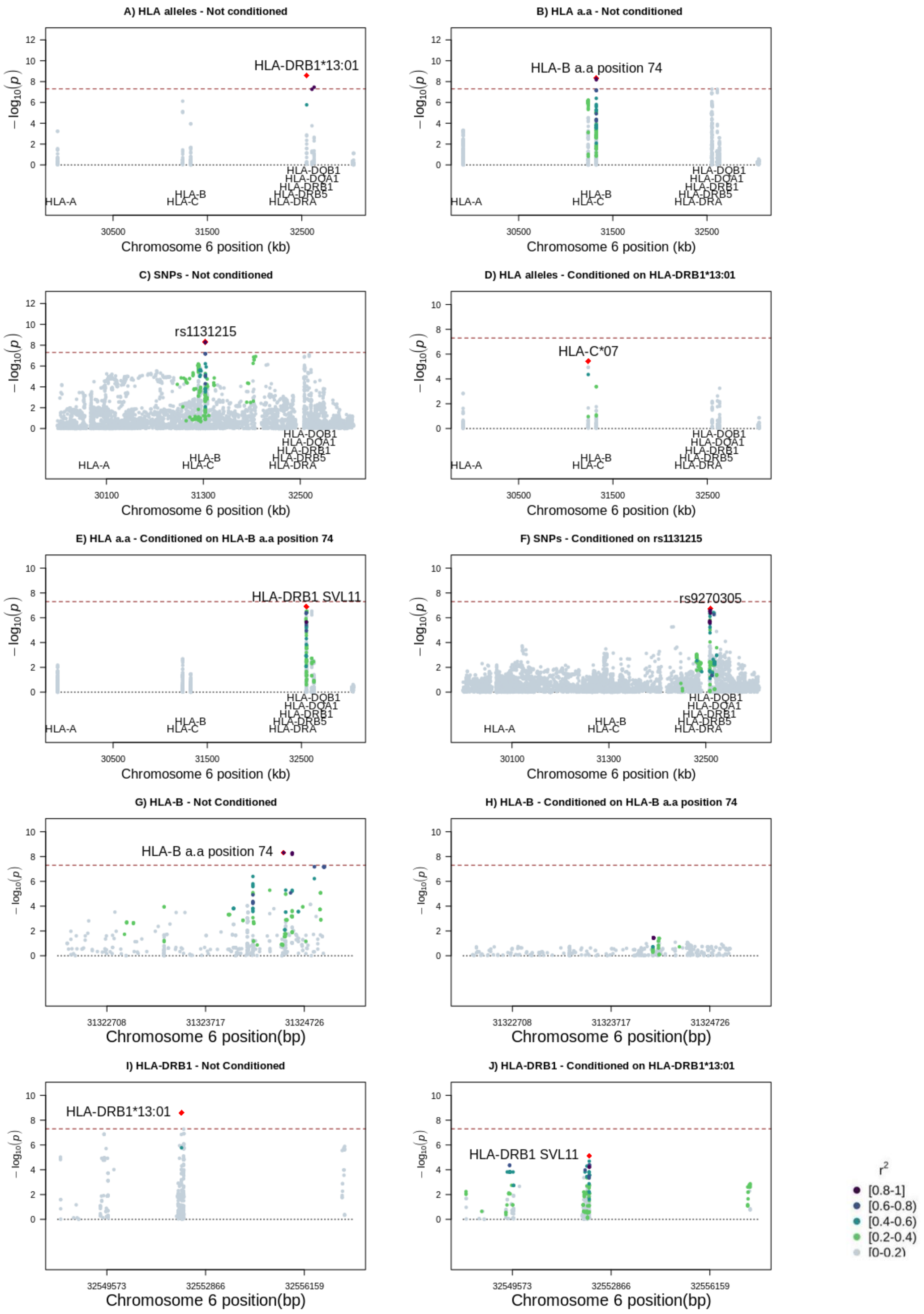
A) HLA alleles - Not conditioned
HLA-DRB1*13:01
B) HLA a.a - Not conditioned
HLA-B a.a position 74
C) SNPs - Not conditioned
rs1131215
D) HLA alleles - Conditioned on HLA-DRB1*13:01
HLA-C*07
E) HLA a.a - Conditioned on HLA-B a.a position 74
HLA-DRB1 SVL11
F) SNPs - Conditioned on rs1131215
rs9270305
G) HLA-B - Not Conditioned
HLA-B a.a position 74
H) HLA-B - Conditioned on HLA-B a.a position 74
I) HLA-DRB1 - Not Conditioned
HLA-DRB1*13:01
J) HLA-DRB1 - Conditioned on HLA-DRB1*13:01
HLA-DRB1 SVL11
HLA-A
HLA-B
HLA-C
HLA-DQB1
HLA-DQA1
HLA-DRB1
HLA-DRB5
HLA-DRA
$-\log_{10}(p)$
Chromosome 6 position (kb)
Chromosome 6 position(bp)
$r^2$
[0.8-1]
[0.6-0.8)
[0.4-0.6)
[0.2-0.4)
[0-0.2)

**Supplementary Figure 12: Regional plots of HLA fine-mapping in MG datasets**. HLA imputation of classical HLA alleles, amino acids (aa) and SNPs was performed using SNP2HLA and Type 1 Diabetes Genetics Consortium data as a reference panel. For the MHC region we used the coordinates in hg19 (chr6:28,477,797-33,448,354) as provided by the Genome Reference Consortium. A) The p-values of classical HLA alleles. B) The p-values of HLA aa. C) The p-values of SNPs in the MHC region. To identify the independently associated loci, conditional analyses were performed including the top hit in the initial association analyses in each type of variant. D) The classical HLA alleles' p-values after conditioning for *HLA-DRB1*13:01*. E) The HLA aa p-values after conditioning for SerValLeu11. F) The SNPs' p-values after conditioning for rs1131215. Panels A-F show *HLA-DRB1* and *HLA-B* as two associated loci. Analyses in each specific locus separately, using all three types of variants in *HLA-B* (panels G-H). indicate that the aa polymorphism at position 74 drives the association signal in *HLA-B*. Similarly, using all three types of variants in *HLA-DRB1* (panels I-J), *HLA-DRB1*13:01* was found to account for the signal in the gene.

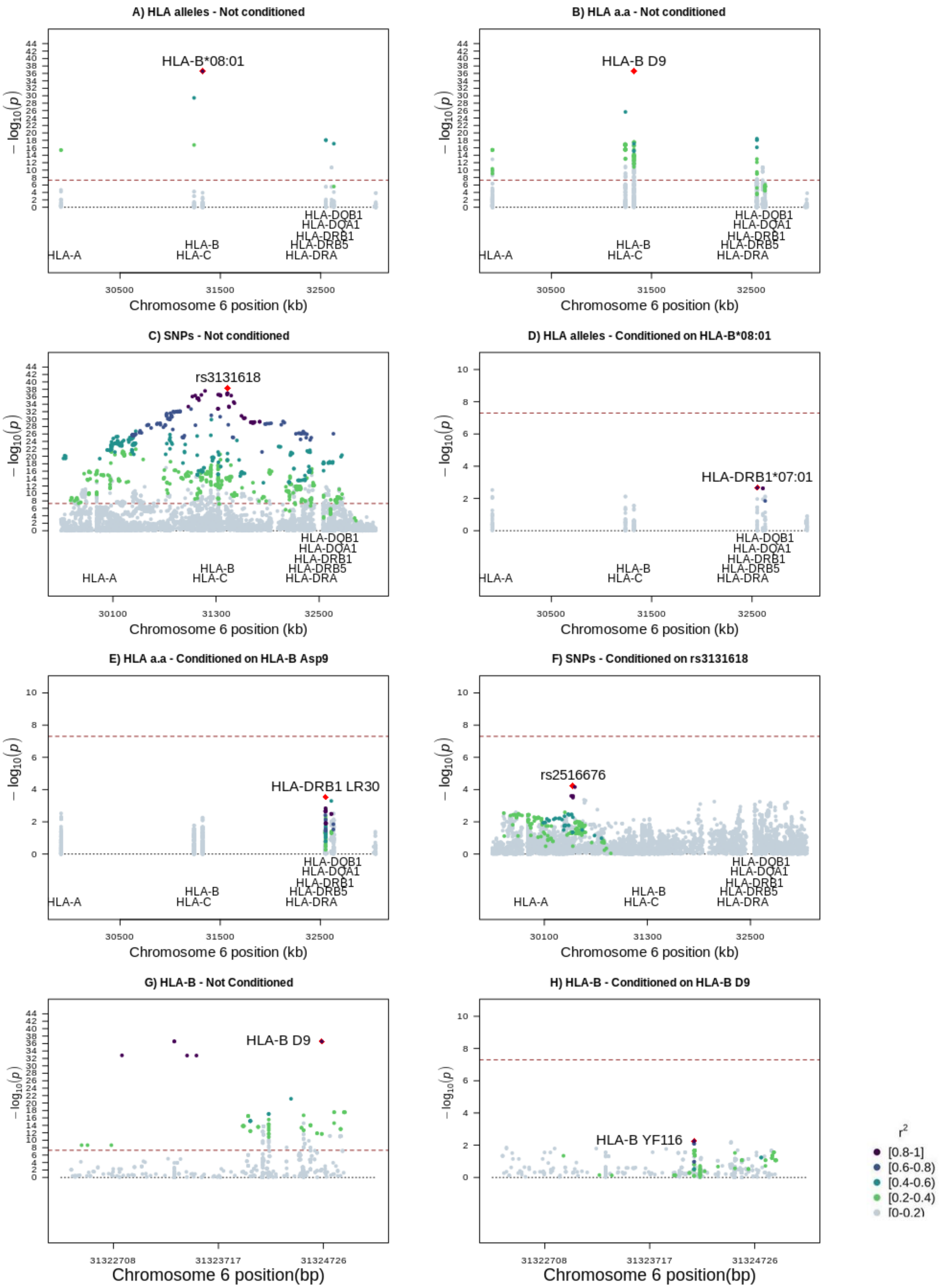

**Supplementary Figure 13: Regional plots of HLA fine-mapping in EOMG datasets**. HLA imputation of classical HLA alleles, amino acids (aa) and SNPs was performed using SNP2HLA and Type 1 Diabetes

Genetics Consortium data as a reference panel. For the MHC region we used the coordinates in hg19 (chr6:28,477,797-33,448,354) as provided by the Genome Reference Consortium. A) The p-values of classical HLA alleles. B) The p-values of HLA aa. C) The p-values of SNPs in the MHC region. To identify the independently associated loci, conditional analyses were performed including the top hit in the initial association analyses in each type of variant. D) The classical HLA alleles' p-values after conditioning for *HLA-B*08:01*. E) The HLA aa p-values after conditioning for Asp9. F) The SNPs' p-values after conditioning for rs2596492. Panels A-H show *HLA-B* as the only independent locus. Analyses using all three types of variants in *HLA-B* (panels G-H) indicate that Asp9 drives the association signal in *HLA-B*.

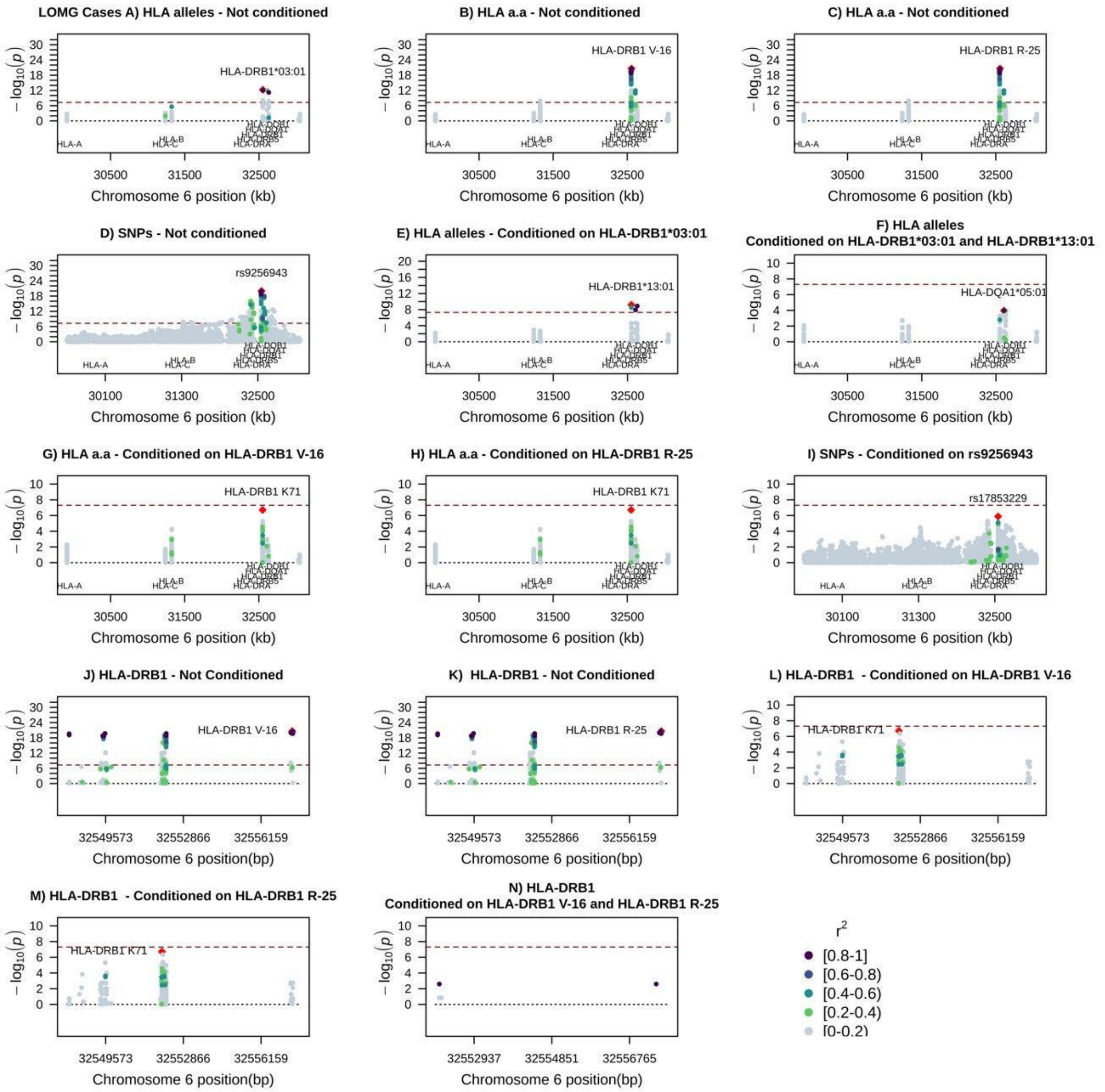

**Supplementary Figure 14: Regional plots of HLA fine-mapping in LOMG datasets.** HLA imputation of classical HLA alleles, amino acids (aa) and SNPs was performed using SNP2HLA and Type 1 Diabetes Genetics Consortium data as a reference panel. For the MHC region we used the coordinates in hg19 (chr6:28,477,797-33,448,354) as provided by the Genome Reference Consortium. A) The p-values of classical HLA alleles. B-C) The p-values of HLA aa. D) The p-values of SNPs in the MHC region. To identify the independently associated loci, conditional analyses were performed including the top hit in the initial association analyses in each type of variant. E) The classical HLA alleles' p-values after conditioning for *HLA-DRB1*03:01*; as *HLA-DRB1*13:01* remained significant, we conditioned for both *HLA-DRB1*03:01* and *HLA-DRB1*13:01*, to check whether there are additional associated loci (Panel F). G-H) The HLA aa p-values after conditioning for Val16 and Arg25, respectively. F) The SNPs' p-values after conditioning for rs9256943. Panels A-H show that *HLA-DRB1* is the only independent locus. Analyses using all three types of variants in *HLA-DRB1* (panels J-N) indicate that aa Val16 and Arg25 have the same effect and account for most of the signal in *HLA-DRB1*. However, when conditioning for both aa, the effect of all variants in the region becomes eliminated.

#### Supplementary Tables

**Supplementary Table 1:** Genetic correlation of MG GWAS with GWAS from other autoimmune disorders. The number of cases and controls in each study is also shown.

**Supplementary Table 2:** LD-independent regions on the MG GWAS meta-analysis, after performing clumping with $p<10^{-5}$ for index SNPs and variants within 3-Mb windows and $r^2>0.1$ with index SNPs.

**Supplementary Table 3:** LD-independent regions on the EOMG GWAS meta-analysis after performing clumping with $p<10^{-5}$ for index SNPs and variants within 3-Mb windows and $r^2>0.1$ with index SNPs.

**Supplementary Table 4:** LD-independent regions on the LOMG GWAS meta-analysis after performing clumping with $p<10^{-5}$ for index SNPs and variants within 3-Mb windows and $r^2>0.1$ with index SNPs.

**Supplementary Table 5**: Comparison of classic HLA alleles associations in previous studies and different onsets. The data show the HLA alleles detected in this study with p-value $<5x10^{-8}$, and the 10 most significantly associated alleles in previous studies. The ORs are shown for each HLA allele, unless not available.
